# Research waste from poor reporting of core methods and results and redundancy in studies of reporting guideline adherence: a meta-research review

**DOI:** 10.1101/2022.12.19.22283669

**Authors:** Tiffany Dal Santo, Danielle B. Rice, Lara S.N. Amiri, Amina Tasleem, Kexin Li, Jill T. Boruff, Marie-Claude Geoffroy, Andrea Benedetti, Brett D. Thombs

**Affiliations:** Lady Davis Institute for Medical Research, Jewish General Hospital, Montreal, Quebec, Canada; Department of Psychiatry, McGill University, Montreal, Quebec, Canada; Department of Psychology, St. Joseph’s Healthcare Hamilton, Ontario, Canada; Department of Psychiatry and Behavioural Neurosciences, McMaster University, Ontario, Canada; Schulich Library of Physical Sciences, Life Sciences, and Engineering, McGill University, Montreal, Quebec, Canada; Department of Educational and Counselling Psychology, McGill University, Montreal, Quebec, Canada; McGill Group for Suicide Studies, Douglas Research Centre, Montreal, Quebec, Canada; Department of Epidemiology, Biostatistics, and Occupational Health, McGill University, Montreal, Quebec, Canada; Department of Medicine, McGill University, Montreal, Quebec, Canada; Respiratory Epidemiology and Clinical Research Unit, McGill University Health Centre, Montréal, Québec, Canada; Biomedical Ethics Unit, McGill University, Montreal, Quebec, Canada; Department of Psychology, McGill University, Montreal, Quebec, Canada

## Abstract

**Objectives:** We investigated meta-research studies that evaluated adherence to prominent reporting guidelines (CONSORT, PRISMA, STARD, STROBE) in health research studies to determine the proportion that (1) provided an explanation for how complex guideline items were rated for adherence and (2) provided results from individual studies reviewed in addition to aggregate results. We also examined the conclusions of each meta-research study to assess redundancy of findings across studies.

**Design:** Cross-sectional meta-research review.

**Data sources:** MEDLINE (Ovid) searched on July 5, 2022.

**Eligibility criteria for selecting studies:** Studies in any language were eligible if they used any version of the CONSORT, PRISMA, STARD, or STROBE reporting guidelines or their extensions to evaluate reporting in at least 10 human health research studies. We excluded studies that modified a reporting guideline or its items or evaluated fewer than half of reporting guideline items. Main outcomes were (1) the proportion of meta-research studies that provided a coding explanation that could be used to replicate the study or verify its results and (2) the proportion that provided individual-level study results in the main text, supplemental materials, or via an internet link.

**Results:** Of 148 included meta-research studies, 14 (10%, 95% confidence interval [CI] 6% to 15%) provided a fully replicable coding explanation, and 49 (33%, 95% CI 26% to 41%) completely reported individual study results. Of 90 studies that classified reporting as adequate or inadequate in the study abstract, 6 (7%, 95% CI 3% to 14%) concluded that reporting was adequate but none of those 6 studies provided information on how items were coded or provided item-level results for included studies.

**Conclusions:** Much of published meta-research on reporting in health research is likely wasteful. Few studies report enough information for verification or replication, and almost all find that reporting in health research studies is suboptimal. These findings highlight the importance of shifting the focus from assessing reporting adequacy to developing, testing, and implementing strategies to improve reporting.

**Funding:** There was no specific funding for this study.

**Protocol:** Posted on the Open Science Framework June 29, 2022 (https://osf.io/gtm4z/).

## INTRODUCTION

Research waste occurs when research is not useful because it addresses questions that are already answered; is too poorly designed to answer important research questions; does not make results easily accessible, or reports results too poorly to evaluate or replicate the research.^1–6^ Meta-research studies are conducted to identify areas where research design, conduct, or reporting could be improved and, thus, reduce research waste.^7^ Just like other health research, however, meta-research can be wasteful if it is too poorly designed or reported to be useful or does not add substantively to knowledge.

Many meta-research studies evaluate whether published studies are adequately reported based on reporting guidelines,^8,9^ such as the Consolidated Standards of Reporting Trials (CONSORT),^10^ Preferred Reporting Items for Systematic Reviews and Meta-Analyses (PRISMA),^11^ Standards for the Reporting of Diagnostic Accuracy Studies (STARD),^12^ or Strengthening the Reporting of Observational Studies in Epidemiology (STROBE).^13^ Reporting guidelines typically include a structured checklist of a minimum set of items that should be reported in studies that use a specific research design along with an explanation document.^14^

How meta-researchers translate reporting guideline items into evaluative ratings of individual studies and the ratings generated for included studies are the core elements of any study on reporting guideline adherence. There are no reporting guidelines for meta-research studies, but many of these studies, including studies on reporting guideline adherence, use methods that closely align with systematic review methods.^15–20^ The PRISMA statement for reporting of systematic reviews and meta-analyses stipulates that methods for collecting and coding data are explicitly defined and that results of each individual study included in a review must be provided.^11^ Many reporting guideline items are complex and include multiple components.^21^ Not defining how these items are translated into ratings in meta-research studies creates risk that coding decisions are not made validly or reliably and poses a barrier to replication. Similarly, the failure to report individual study-level results, at least in supplemental material or via a weblink, poses a barrier to reviewing and verifying aggregate findings and does not allow users to identify studies with specific findings that may be of interest.

The objective of our study was to evaluate meta-research studies that reviewed reporting in health research studies using the CONSORT,^10^ PRISMA^11^, STARD,^12^ or STROBE^13^ reporting guidelines or one of their extensions to determine the proportion that (1) provided an explanation for how reporting guideline items were translated into adherence ratings with enough information to be replicable and (2) provided results from each individual study included in their report. Additionally, we evaluated the conclusions from the abstract of each study to assess the degree to which meta-research studies on reporting are likely generating new knowledge versus addressing a question for which the answer may already be known.

## METHODS

We conducted a cross-sectional evaluation of recently published meta-research studies that evaluated adequacy of health research study reporting based on any version of the CONSORT,^10^ PRISMA,^11^ STARD,^12^ or STROBE^13^ reporting guidelines or their extensions. We posted our study protocol on the Open Science Framework (https://osf.io/gtm4z/) prior to initiation. Although there are currently no reporting guidelines for meta-research studies, we have reported the present study consistent with applicable PRISMA items as these most closely align with our study design.

### Eligibility

Studies published in any language were eligible if they used any version of the CONSORT,^10^ PRISMA,^11^ STARD,^12^ or STROBE^13^ reporting guidelines or their extensions (e.g., CONSORT-ROUTINE,^22^ PRISMA-DTA,^23^ STROBE-MR^24^) to evaluate reporting in human health research publications. To select reporting guidelines for inclusion in our study, we reviewed citations of all EQUATOR guideline publications^21^ and found that these guidelines were by far the most highly cited. To facilitate searching, included studies must have mentioned the name of an eligible guideline in their abstract. Studies that evaluated reporting using multiple reporting guidelines were eligible if at least one of the guidelines was eligible. Studies that investigated reporting as one of multiple research questions or that assessed reporting as part of another research question were eligible. We excluded studies that modified items in an otherwise eligible reporting guideline checklist, added items to an eligible checklist, or evaluated fewer than half of items in an eligible checklist as this could create small subsets of items that might have a different level of coding complexity compared to the full list of checklist items. We excluded studies that evaluated fewer than 10 publications.

### Search and Study Selection Method

We searched MEDLINE (ALL) via Ovid for potentially eligible studies using the search strategy: (((quality or complete* or adequat* or transparen*) adj3 reporting) AND (CONSORT* or PRISMA* or STROBE* or STARD* or “Consolidated Standards of Reporting Trials” or “Preferred Reporting Items for Systematic Reviews” or “Standards for Reporting Diagnostic accuracy studies” or “Strengthening the Reporting of Observational Studies in Epidemiology”)).tw,kf. The principal investigator (BDT) worked with an experienced health sciences librarian (JTB) to develop the search. The search was then run by a trained research assistant (KL) on July 5, 2022. To include the most recently published meta-research studies and reflect current practices as best as possible, we reviewed citations identified in the search in reverse chronological order based on their PubMed Unique Identifier until we obtained our targeted sample size of eligible studies. Citations were uploaded to DistillerSR (Evidence Partners, Ottawa, Canada). Two reviewers (TDS, LSNA) independently assessed study eligibility at the title and abstract level. If either reviewer believed a study to be potentially eligible, it moved on to full-text review, where two reviewers (TDS, LSNA, AT) independently assessed eligibility. Discrepancies at the full-text level were resolved by consensus between reviewers with a third reviewer (BDT) consulted as necessary. Supplementary Material 1 includes coding criteria for determining eligibility at the title and abstract and full-text levels.

### Sample Size Calculation

A preliminary pre-study review of likely eligible articles published in the 5 years prior to July 2022 found that few studies provided coding definitions or reported individual study results. We therefore hypothesized that the proportion of included articles that provided either would be small. Thus, we set our sample size to have a 95% confidence interval (CI) width of 15% around a percentage reporting of 33%. Based on CIs calculated using the method of Agresti and Coull,^25^ we sought to obtain 148 studies.

### Data Extraction

For each eligible meta-research study, data were extracted in DistillerSR by a single reviewer (TDS, LSNA) and validated by a second reviewer (TDS, LSNA, AT) using the DistillerSR Quality Control function. Discrepancies were resolved by consensus between reviewers with a third reviewer (BDT) consulted as necessary. The data extraction form can be found in Supplementary Material 2. Reviewers extracted (1) publication characteristics (first author last name; publication year; journal and 2021 impact factor); (2) corresponding author country; (3) research question (reporting was the only research question; reporting was the main research question and there were other non-reporting questions; there were multiple research questions, including reporting and non-reporting questions, and the main one is unclear; the main research question was not reporting, but an eligible reporting analysis was conducted) (4) reporting guideline(s) evaluated; (5) number of publications included in the study; (6) main eligibility criteria of included publications (by reporting guideline, study design, field of research, patient population, intervention type, journal, other); (7) number and independence of raters and the rating method used (e.g., yes/no, fully/partially/not reported); and (8) conclusion about reporting adequacy. We used publication abstracts to extract conclusions as these are the most read, and in many cases, the only part of an article that is read.^26,27^ If a study’s supplementary material was not accessible via the publishing journal’s website, we contacted the corresponding author and the journal editorial manager or editor-in-chief to request access. We sent up to 2 follow-up emails to corresponding authors and journal staff until access was obtained; if we did not receive a response, we coded the study based on available information because no study user would have access to the supplementary material.

To answer our main research questions, reviewers extracted (1) whether the authors provided coding explanations for translating items into ratings with enough information to be replicated and (2) if the authors provided results for each individual study included in their report. Authors may have provided coding explanations and individual study results in the main text or table of their study, in supplementary material, or via an internet link. Coding explanations must have specifically reported which parts of each item were required for the item to be used in evaluating reporting adequacy. For individual study results, we coded whether authors reported results for each item for all studies, reported results partially (e.g., an overall score but not item ratings for each study), or did not report individual study results. Our coding manual for evaluating coding explanations and reporting of individual study results are found in Supplementary Material 3 and 4.

### Analysis

We calculated the (1) proportion of meta-research studies that provided a coding guide for translating reporting guideline items into ratings with enough information to be replicated and (2) proportion that provided results for each individual study included in their report. All proportions are presented with 95% CIs using the method of Agresti and Coull.^25^ We also presented results by subgroups defined by corresponding author country, 2021 journal impact factor, reporting guideline evaluated (CONSORT, PRISMA, STROBE, STARD), and research question (reporting is part of the main research question; reporting is not part of the main research question). When presenting outcomes by subgroups, we included guideline extensions (e.g., CONSORT-ROUTINE) with the main guideline (e.g., CONSORT). Groupings (e.g., impact factor levels) were established based on available frequency data prior to examining reporting results. We did not conduct statistical tests to compare subgroups because our study was not designed or powered for that purpose.

### Patient and Public Involvment

Patients and members of the public were not involved in the design, conduct, reporting, or dissemination plans of our research.

## RESULTS

### Search Results and Included Study Characteristics

Our search yielded 1,698 unique titles and abstracts. We reviewed these in reverse chronological order until we obtained 148 included studies. During this process we reviewed 418 titles and abstracts. We excluded 182 studies at the title and abstract level, and 88 studies at the full-text level (Figure 1).

**Figure 1.**
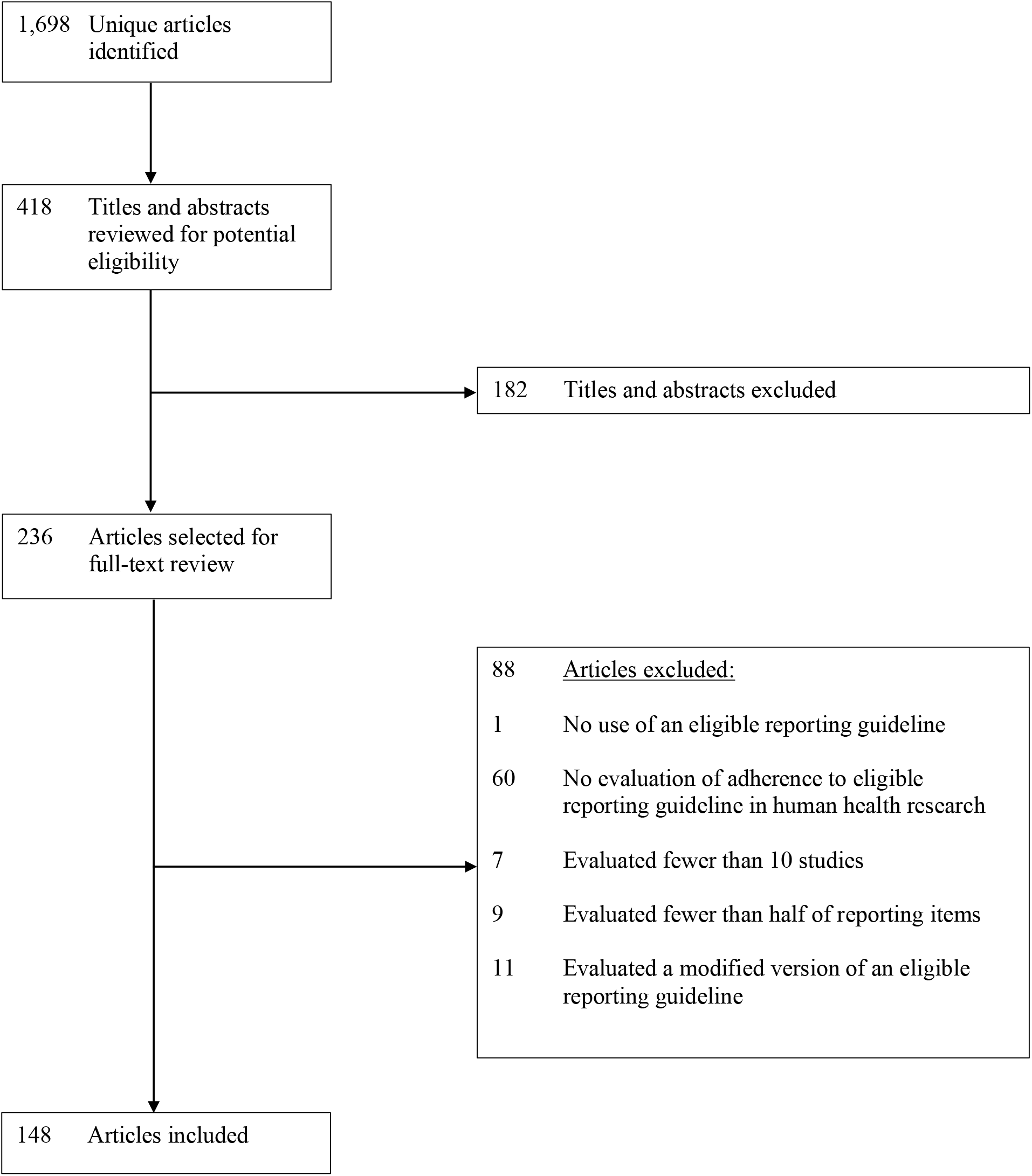
PRISMA flow diagram.

The 148 included studies were initially listed on PubMed between August 14, 2020 and June 30, 2022. They assessed between 10 and 2,844 studies (median = 52; interquartile range = 24 to 120). As shown in Table 1, included studies were from China (N = 51; 34%), the United States (N = 27; 18%), the United Kingdom (N = 9; 6%), Canada (N = 8; 5%), and 22 other countries that had 1 to 5 included studies (N = 53; 36%). Most studies assessed adherence to the CONSORT (N = 61; 41%) or PRISMA (N = 59; 40%) reporting guidelines or their extensions. Reporting was the only research question in 46 (31%) studies, the main question among multiple questions in 13 (9%) studies, one of multiple questions with no clear primary question in 65 (44%) studies, and not part of the main question in 24 (16%) studies. Most studies 103 (70%) came from journals with impact factor > 2.9. See Supplementary Material 5 for individual study characteristics.

**Table 1:** Study characteristics (N = 148)

| **Study Characteristics** | **N (%)** |
| --- | --- |
| **Year Published** |  |
| 2020 | 21 (14) |
| 2021 | 60 (41) |
| 2022 | 56 (38) |
| Online only | 11 (7) |
| **Country** |  |
| Canada | 8 (5) |
| China | 51 (34) |
| United Kingdom | 9 (6) |
| United States | 27 (18) |
| Other (all with $\leq 5$ studies)<sup>a</sup> | 53 (36) |
| **Journal Impact Factor<sup>b</sup>** |  |
| Impact factor $\leq 2.9$ | 45 (30) |
| Impact factor $> 2.9$ | 103 (70) |
| **Included Study Eligibility Criteria<sup>c</sup>** |  |
| Study design | 137 (93) |
| Patient population | 68 (46) |
| Intervention type | 65 (44) |
| Journal | 17 (11) |
| Included in specified guidelines | 13 (9) |
| Field of research | 13 (9) |
| Other<sup>d</sup> | 6 (4) |
| **Research Question** |  |
| Reporting is only research question | 46 (31) |
| Reporting is main of multiple research questions | 13 (9) |
| Multiple research questions with main unclear | 65 (44) |
| Main research question not reporting | 24 (16) |
| **Reporting Guideline<sup>e</sup>** |  |
| CONSORT | 61 (41) |
| PRISMA | 59 (40) |
| STARD | 10 (7) |
| STROBE | 18 (12) |
| **Number of Included Publications Reviewed** |  |
| $\leq 50$ | 72 (49) |
| $>50$ | 76 (51) |

<sup>a</sup>Australia (3); Brazil (3); Chile (1); Croatia (1); France (3); Germany (4); Greece (2); India (3); Iran (2); Ireland (2); Italy (3); Korea (4); Macao (2); Mexico (1); Portugal (1); Qatar (2); Saudi Arabia (2); South Africa (1); South Korea (4); Spain (3); Switzerland (1); the Netherlands (5). <sup>b</sup>Journals for which we could not find an impact factor were coded as 0. <sup>c</sup>Included reviews could be counted in more than one category. <sup>d</sup>Studies reviewed included a specific questionnaire, were on acceptability of a specific intervention, were abstracts submitted to specific conferences, or were studies that used a specific database. <sup>e</sup>Including extensions to specified reporting guidelines.

Of the 148 included studies, 3 studies (2%, 95% CI 1% to 6%) used 1 rater, 10 (7%, 95% CI 4% to 12%) used 1 rater with validation from a second rater, 113 (76%, 95% CI 69% to 83%) used 2 or more independent raters, 9 (6%, 95% CI 3% to 11%) used 2 or more raters but did not state whether they were independent, 3 (2%, 95% CI 1% to 6%) used other methods, and 10 (7%, 95% CI 4% to 12%) did not report how many raters were used. For classifying adherence to reporting checklist items, 66 studies (45%, 95% CI 37% to 53%) classified items dichotomously, 61 (41%, 95% CI 34% to 49%) used a multi-level approach (e.g., coded item as “fully reported”, “partially reported”, or “not reported”), 2 (1%, 95% CI 0% to 5%) classified some items dichotomously and others with a multi-level approach, and 19 (13%, 95% CI 8% to 19%) did not report how they classified items. See Supplementary Material 6.

### Main Outcomes

Of the 148 studies, 14 (10%, 95% CI 6% to 15%) provided a fully replicable coding explanation, 5 (3%, 95% CI 2% to 8%) provided a partially replicable coding explanation, and 129 (87%, 95% CI 81% to 92%) did not provide enough information to know how coding decisions had been made (see Table 2). Forty-nine studies (33%, 95% CI 26% to 41%) completely reported individual study results, 26 (18%, 95% CI 12% to 25%) reported partial results for all studies, 3 (2%, 95% CI 1% to 6%) reported results for some studies but not others, and 70 (47%, 95% CI 39% to 55%) did not provide any individual study results (see Table 3).

**Table 2:** Number and percent of studies that provided fully or partially replicable coding explanations or did not provide coding explanations for overall sample (N = 148) and subgroups.

| Subgroups | N<br>% (95%CI) |  |  |
| --- | --- | --- | --- |
|  | Fully Replicable | Partially Replicable | Not Replicable |
| **All** | 14<br>10% (6%, 15%) | 5<br>3% (2%, 8%) | 129<br>87% (81%, 92%) |
| **Country** |  |  |  |
| Canada | 3<br>38% (14%, 69%) | 2<br>25% (7%, 59%) | 3<br>38% (14%, 69%) |
| China | 3<br>6% (2%, 16%) | 0<br>0% (0%, 7%) | 48<br>94% (84%, 98%) |
| United Kingdom | 1<br>11% (2%, 44%) | 1<br>11% (2%, 44%) | 7<br>78% (45%, 94%) |
| United States | 2<br>7% (2%, 23%) | 0<br>0% (0%, 13%) | 25<br>93% (77%, 98%) |
| Other | 5<br>9% (4%, 20%) | 2<br>4% (1%, 13%) | 46<br>87% (75%, 94%) |
| **Journal Impact Factor** |  |  |  |
| Impact factor $\leq 2.9$ | 1<br>2% (0%, 12%) | 0<br>0% (0%, 8%) | 44<br>98% (88%, 100%) |
| Impact factor $> 2.9$ | 13<br>13% (8%, 20%) | 5<br>5% (2%, 11%) | 85<br>83% (74%, 89%) |
| **Reporting Guideline** |  |  |  |
| CONSORT & extensions | 9<br>15% (8%, 26%) | 3<br>5% (2%, 14%) | 49<br>80% (69%, 88%) |
| PRISMA & extensions | 1<br>2% (0%, 9%) | 1<br>2% (0%, 9%) | 57<br>97% (89%, 99%) |
| STARD & extensions | 2<br>20% (6%, 51%) | 1<br>10% (2%, 40%) | 7<br>70% (40%, 89%) |
| STROBE & extensions | 2<br>11% (3%, 33%) | 0<br>0% (0%, 18%) | 16<br>89% (67%, 97%) |
| **Research Question** |  |  |  |
| Reporting is the only research question or there are multiple research questions and the main one is reporting or not defined | 14<br>11% (7%, 18%) | 4<br>3% (1%, 8%) | 106<br>86% (78%, 91%) |
| Reporting is not the main research question | 0<br>0% (0%, 14%) | 1<br>4% (1%, 20%) | 23<br>96% (80%, 99%) |

**Table 3:** Level of reporting of included study results for overall sample (N = 148) and subgroups.

| Subgroups | N<br>% (95%CI) |  |  |  |
| --- | --- | --- | --- | --- |
|  | Completely Reported | Partially Reported –<br>All Studies | Partially Reported –<br>Some Studies | Not Reported |
| **All** | 49<br>33 % (26%, 41%) | 26<br>18% (12%, 25%) | 3<br>2% (1%, 6%) | 70<br>47 % (39%, 55%) |
| **Country** |  |  |  |  |
| Canada | 2<br>25% (7%, 59%) | 0<br>0% (0%, 32%) | 0<br>0% (0%, 32%) | 6<br>75% (41%, 93%) |
| China | 26<br>51% (38%, 64%) | 2<br>4% (1%, 13%) | 0<br>0% (0%, 7%) | 23<br>45% (32%, 59%) |
| United Kingdom | 4<br>44% (19%, 73%) | 2<br>22% (6%, 55%) | 0<br>0% (0%, 30%) | 3<br>33% (12%, 65%) |
| United States | 3<br>11% (4%, 28%) | 13<br>48% (31%, 66%) | 1<br>4% (1%, 18%) | 10<br>37% (22%, 56%) |
| Other | 14<br>26% (16%, 40%) | 9<br>17% (9%, 29%) | 2<br>4% (1%, 13%) | 28<br>53% (40%, 66%) |
| **Journal Impact Factor** |  |  |  |  |
| Impact factor ≤ 2.9 | 16<br>36% (23%, 50%) | 9<br>20% (11%, 34%) | 1<br>2% (0%, 12%) | 19<br>42% (29%, 57%) |
| Impact factor > 2.9 | 33<br>32% (24%, 42%) | 17<br>17% (11%, 25%) | 2<br>2% (1%, 7%) | 51<br>50% (40%, 59%) |
| **Reporting Guideline** |  |  |  |  |
| CONSORT & extensions | 14<br>23% (14%, 35%) | 6<br>10% (5%, 20%) | 0<br>0% (0%, 6%) | 41<br>67% (55%, 78%) |
| PRISMA & extensions | 25<br>42% (31%, 55%) | 15<br>25% (16%, 38%) | 1<br>2% (0%, 9%) | 18<br>31% (20%, 43%) |
| STARD & extensions | 3<br>30% (11%, 60%) | 1<br>10% (2%, 40%) | 1<br>10% (2%, 40%) | 5<br>50% (24%, 76%) |
| STROBE & extensions | 7<br>39% (20%, 61%) | 4<br>22% (9%, 45%) | 1<br>6% (1%, 26%) | 6<br>33% (16%, 56%) |
| **Research Question** |  |  |  |  |
| Reporting is the only research question or there are multiple research questions and the main one is reporting or not defined | 35<br>28% (21%, 37%) | 22<br>18% (12%, 25%) | 2<br>2% (0%, 6%) | 65<br>52% (44%, 61%) |
| Reporting is not the main research question | 14<br>58% (39%, 76%) | 4<br>17% (7%, 36%) | 1<br>4% (1%, 20%) | 5<br>21% (9%, 41%) |

Only 4 (3%, 95% CI 1% to 7%) studies provided both fully replicable coding explanations and completely reported individual study results.

One hundred and twenty-two studies mentioned reporting in their abstract conclusion, and 90 of these classified reporting as either adequate or inadequate. Of these 90 studies, 6 (7%, 95% CI 3% to 14%) concluded that reporting was adequate, 29 (32%, 95% CI 24% to 42%) implicitly concluded that reporting was inadequate, and 55 (61%, 95% CI 51% to 71%) explicitly concluded that reporting was inadequate. Of the 6 studies that concluded that reporting was adequate, none provided any explanation of how items were coded, and none provided item-level results for individual studies. The 4 studies with fully replicable coding explanations and complete individual study results all concluded that reporting of individual studies was inadequate. See Table 4. Outcomes for all individual meta-research studies we reviewed are shown in Supplementary Material 7.

**Table 4:**
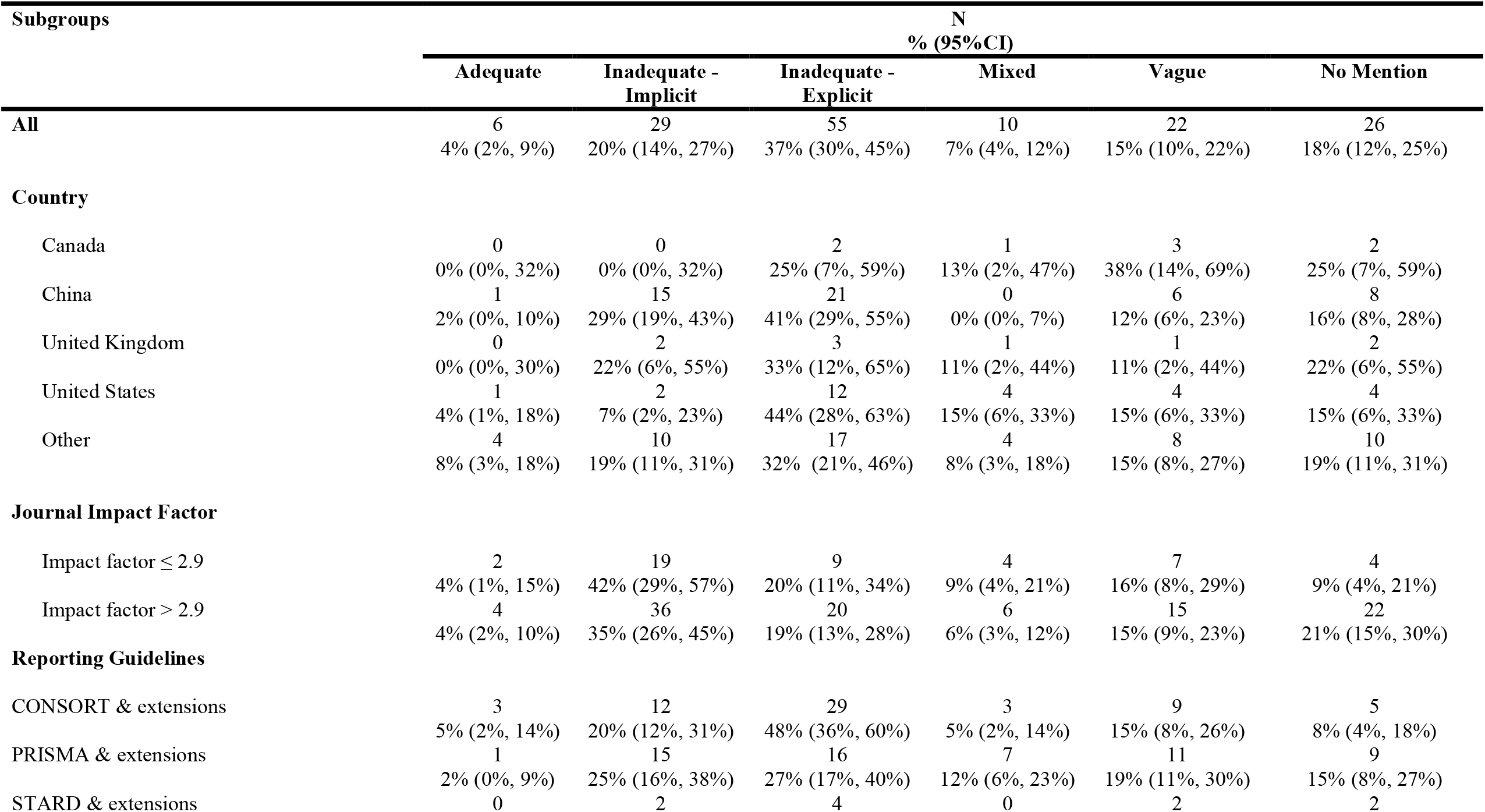
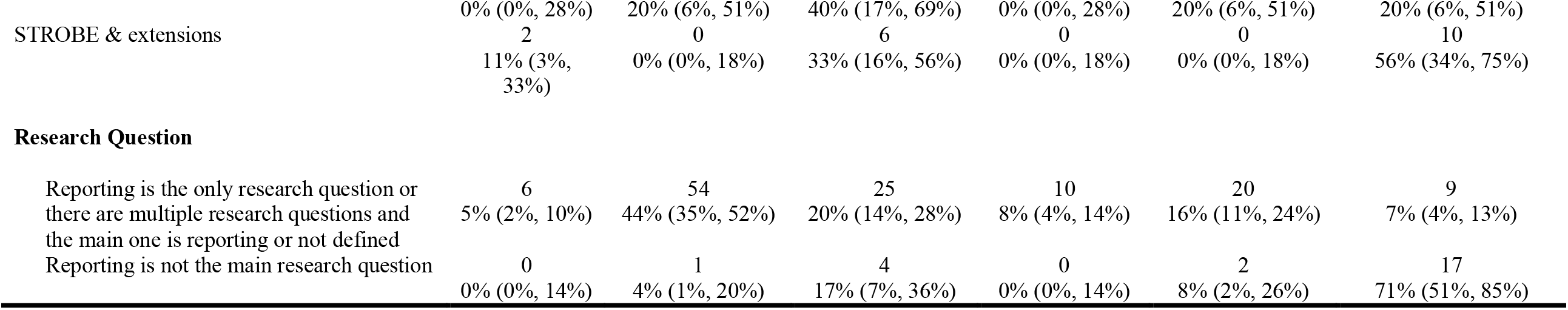
Conclusions in abstracts of included studies on research reporting for overall sample (N = 148) and subgroups.

As shown in Tables 2, 3 and 4, most subgroup results did not differ substantively from overall conclusions, excluding subgroups with very small numbers of meta-research studies (e.g., < 10 studies). One exception was for subgroups by the research question. Of the 124 studies where reporting was part of the main research questions, 35 (28%, 95% CI 21% to 37%) completely reported individual study results, whereas 14 of 24 studies (58%, 95% CI 39% to 76%) where reporting was not the main research question reported individual study results. These 24 studies were mostly systematic reviews with clinical questions that assessed reporting as part of their evaluation of included studies.

## DISCUSSION

We examined 148 studies that evaluated reporting guideline adherence across areas of health research and were initially listed in PubMed between August 2020 and June 2022. Of the 148 studies, only 10% provided enough information on how individual checklist items were rated to allow for verification of how study methods had been applied or for others to replicate them. Only 33% reported results for all studies they evaluated, at least in supplementary material or via an internet link, so that results could be verified. We did not identify any substantive differences by subgroups such as country or impact factor of the journal where studies were published. Of 90 studies that classified reporting as adequate or inadequate in their abstracts, 7% concluded that reporting was adequate; however, none of the studies that described reporting as adequate provided an explanation of how they coded items or provided item-level results for individual studies. Only 3% of included meta-research studies provided both fully replicable coding explanations and complete individual study results, and all of those studies concluded that reporting was inadequate.

No previous studies have examined the degree to which meta-research studies on reporting guideline adherence adequately report key aspects of their own studies. Given that meta-research is done to scrutinize research methodology and conduct and to promote methods that support the validity and integrity of science,^28^ an implicit assumption is that meta-research studies themselves are rigorously conducted and reported. Our study shows that this is often not the case. Dozens of reviews of studies on health research reporting have underlined that reporting is consistently poor.^29^ Our findings show that many of the meta-research studies that have led to those conclusions are themselves poorly reported.

Good research asks important questions and uses methods that allow us to be confident in its conclusions.^30^ Large numbers of studies are being done on reporting adherence, virtually all finding the same thing, but few of them are well-enough conducted and reported for more nuanced conclusions to be drawn with confidence. Researchers who are considering initiating a study on adherence to reporting guidelines and editors who must decide whether to publish such studies should be able to clearly articulate the specific problem they are addressing and how they might add to what we already know about the state of research reporting. For example, evaluating reporting to understand the influence of new or modified reporting guidelines or to assess the effects of interventions designed to improve reporting would likely be justified.

Simply documenting poor reporting guideline adherence in one more sub-specialty area would likely not be useful. Authors of any studies that do evaluate reporting should clearly describe how reporting was evaluated and should provide study-level information so that others can evaluate and validate their findings. Reporting guidelines for meta-research studies do not yet exist, but a protocol for such guidelines has been published.^16^ The authors of these proposed guidelines should ensure that meta-research studies on reporting, in addition to other important items, address the reporting gaps we have identified here.

Rather than more studies on the poor quality of reporting in health care research, interventions that help researchers, peer reviewers, and journal editors improve reporting are needed. A 2019 scoping review identified 31 interventions created to improve reporting guideline adherence but only 11 that had been evaluated in any way.^31^ Strategies varied on what step of the writing or publishing process they targeted, but most have been aimed at improving adherence at the journal level, such as editorial endorsement of specific reporting guidelines, instructions to authors that recommend or require adherence to reporting guidelines, or requiring authors to submit a completed reporting checklist. The scoping review found only 4 randomised trials of interventions to enhance adherence, but the only one that showed a statistically significant effect of the intervention was the Consort-based WEB (COBWEB) tool, that supports adherence at the writing stage of the manuscript.^32^ The tool divides CONSORT items into bullet points and emphasizes key reporting elements that need to be reported for the main CONSORT checklist and selected extensions.^32^ In the trial of the COBWEB tool, which included 41 participants, the global score for completeness of reporting (0-10 scale) was 2.1 points higher (95% CI 1.5 to 2.7) in 123 CONSORT domains drafted with the tool compared to 123 domains drafted without the use of the tool.^32^ Another intervention, which was published after the search period of the scoping review, in which a journal required authors to incorporate section headings that reflected CONSORT items into their manuscripts, also improved reporting.^33^ Overall, however, there are few interventions that have been tested in randomised trials and found to be effective. Resources should be allocated to developing, testing, and disseminating effective interventions that address different aspects of the complex factors that contribute to how well research is reported.^31^

### Strengths and Limitations

Strengths of our study include that it is the first study, to the best of our knowledge, to assess reporting in meta-research studies on research reporting and, thus, addresses an important and largely unrecognized problem. We developed and posted a protocol prior to initiating the study, and we have provided all our coding manuals and individual study results in supplementary material. We included a large sample size of the most recently published studies based on an a priori power analysis.

There are some limitations that also need to be considered. First, we only searched MEDLINE, which could have led us to miss potentially eligible studies, although we found that reporting was poor, and it is unlikely that health research studies in other databases, but not MEDLINE, would have been more completely reported. Second, we included meta-research studies that assessed adherence to 4 EQUATOR reporting guidelines based on how often they have been cited, but we did not assess other reporting guidelines. We do not believe that including other reporting guidelines would have influenced results substantively considering that we assessed reporting in the meta-research studies themselves and not reporting levels of studies that used those reporting guidelines.

## CONCLUSIONS

To conclude, we found that out of the 148 studies we assessed, 10% provided a fully replicable coding explanation, 33% completely reported individual study results, and 7% of those that categorized reporting as being adequate or inadequate concluded that adherence to reporting guidelines was adequate, though none of the studies that rated reporting as adequate were themselves well reported. Meta-research is done to reduce research waste by improving how research is performed, communicated, and used,^28^ but our study shows that meta-research on reporting may be a significant contributor to waste. Most recent studies on reporting guideline adherence do not appear to have added meaningfully to what we know about the problem of research reporting. Poor reporting of key elements in most of these studies does now allow us to draw conclusions beyond that overall reporting continues to be sub-optimal, such as understanding where reporting gaps are most salient or how to address them. New studies on adherence should only be conducted if there is a specific and justified rationale to address a well-defined, non-redundant research question, such as whether implementation of an intervention changes reporting practices. Rather than more research on poor reporting in yet another sub-specialty area, research is needed that develops effective interventions to improve reporting, tests them in randomised trials, and disseminates them via support and training tools.

## Data Availability

All data used in the study are available in the manuscript and its tables or supplementary files.

## Contributions

TDS, DBR, MCG, AB, and BDT were responsible for the study conception and design. JTB and BDT were responsible for the design of the database search. KL carried out the search. TDS, LSNA, AT, and BDT, contributed to data extraction, coding, and evaluation of included studies. TDS conducted the analyses. TDS drafted the manuscript, and DBR, LSNA, AT, KL, JTB, MCG, AB, and BDT provided critical review and approved the final manuscript.

## Copyright for Authors

The Corresponding Author has the right to grant on behalf of all authors and does grant on behalf of all authors, to the Publishers and its licensees in perpetuity, in all forms, formats and media (whether known now or created in the future), to i) publish, reproduce, distribute, display and store the Contribution, ii) translate the Contribution into other languages, create adaptations, reprints, include within collections and create summaries, extracts and/or, abstracts of the Contribution, iii) create any other derivative work(s) based on the Contribution, iv) to exploit all subsidiary rights in the Contribution, v) the inclusion of electronic links from the Contribution to third party material where-ever it may be located; and, vi) licence any third party to do any or all of the above.

The Corresponding Author has the right to grant on behalf of all authors and does grant on behalf of all authors, an exclusive licence (or non-exclusive for government employees) on a worldwide basis to the BMJ Publishing Group Ltd to permit this article (if accepted) to be published in BMJ editions and any other BMJPGL products and sublicences such use and exploit all subsidiary rights, as set out in our licence.

## Ethical Approval

As this study was a meta-research study of published results, it did not require ethical approval.

## Transparency Declaration

The manuscript’s guarantor affirms that the manuscript is an honest, accurate, and transparent account of the study being reported; that no important aspects of the study have been omitted; and that any discrepancies from the study as planned (and, if relevant, registered) have been explained.

## Data Sharing

All data used in the study are available in the manuscript and its tables or supplementary files.

## Dissemination to study participants or patient communities

There are no plans to disseminate the results of the research directly to a relevant patient community.

## Provenance and peer review

Not commissioned; externally peer reviewed.

## CC BY licence

The default licence, a CC BY NC licence, is needed.

## What is already known on this topic

- Many meta-research studies have reported that most health research does not adhere to reporting guidelines.
- Meta-research studies are presumably conducted rigorously and fill an important role of reducing waste by identifying areas where we can improve research practice; no studies have evaluated the degree to which meta-research studies on reporting are themselves well-enough reported and add meaningfully to knowledge to fill this role.

## What this study adds

- We reviewed 148 meta-research studies published in 2020 to 2022 that evaluated reporting guideline adherence in human health research.
- We found that only 10% of meta-research studies included enough information on their data coding methods to verify results or replicate the studies, only 33% provided results for their coding of individual included studies, and almost all reached the same conclusion that reporting is not adequate.
- Our findings demonstrate that much meta-research on reporting guideline adherence likely contributes to research waste due to poor reporting and redundancy, and we need to shift our focus to developing, testing, and disseminating effective strategies to improve reporting.

