## Supplementary File for "Research waste from poor reporting of core methods and results and redundancy in studies of reporting guideline adherence: a meta-research review"

**Supplementary Material: Table of Contents**

**Supplementary Material 1.** Title and abstract and full-text coding guides

**Supplementary Material 2.** Data extraction form

**Supplementary Material 3.** Coding guide for provision of an adequate coding explanation

**Supplementary Material 4.** Coding guide for assessing reporting of included study results

**Supplementary Material 5.** Characteristics of included studies

**Supplementary Material 6.** Rating method and number of raters for overall sample and subgroups

**Supplementary Material 7.** Outcomes for all included studies

**Supplementary Material 1: Title and Abstract and Full-text Coding Guides**

TITLE AND ABSTRACT INCLUSION AND EXCLUSION CRITERIA CODING GUIDE:

**No: no use of eligible reporting guideline.** Exclude if the title or abstract of the article does not mention a version of any eligible reporting guideline (CONSORT, PRISMA, STARD, STROBE), or any eligible extension (e.g., CONSORT-ROUTINE, PRISMA-DTA, STROBE-MR).

**No: no evaluation of adherence to eligible reporting guideline in human health research.** Exclude if it is clear from the title or abstract that the article does not evaluate reporting guideline adherence in a human health research publication.

**No: evaluates single or small set of studies.** Exclude if it is clear from the title or abstract that the article evaluates only a single study or small (< 10) set of studies.

**No: evaluates fewer than half of reporting items.** Exclude if it is clear from the title or abstract that the article evaluates fewer than half of the items in an eligible checklist.

**No: evaluates a modified version of an eligible reporting guideline.** Exclude if it is clear from the title or abstract that the article evaluates a reporting guideline that has been modified.

**Yes: study eligible for inclusion in full-text review.**

FULL-TEXT INCLUSION AND EXCLUSION CRITERIA CODING GUIDE:

**No: no use of eligible reporting guideline.** Exclude if the article does not mention a version of any eligible reporting guideline (CONSORT, PRISMA, STARD, STROBE), or any eligible extension (e.g., CONSORT-ROUTINE, PRISMA-DTA, STROBE-MR).

**No: no evaluation of adherence to eligible reporting guideline in human health research.** Exclude if the article does not evaluate reporting guideline adherence in a human health research publication. Exclude if not all included studies are in human health research.

**No: evaluates single or small set of studies.** Exclude if the article evaluates only a single study or small (< 10) set of studies.

**No: evaluates fewer than half of reporting items.** Exclude if the article evaluates fewer than half of the items in an eligible checklist.

**No: evaluates a modified version of an eligible reporting guideline.** Exclude if the article evaluates a reporting guideline that has been modified.

**Yes: study eligible for inclusion in the meta-research study.**

**Supplementary Material 2: Data Extraction Form**

**1) First Author, Last Name** [textbox]

**2) Year Published** [dropdown]

- Online only
- 2022
- 2021
- 2020
- 2019
- 2018
- 2017

**3) Full Journal Name** [textbox]

**4) 2021 Journal Impact Factor (To be extracted post hoc)** [note]

**5) Country of Corresponding Author** [textbox]

Note: Don’t use abbreviations

**6) Research Question** [dropdown]

o	Reporting is the only research question (including factors associated with reporting)

o	Reporting is the main research question (including factors associated with reporting) and there are other non-reporting questions

o	There are multiple research questions, including reporting and non-reporting questions, and the main one is unclear

o	The main research question is not reporting, but an eligible reporting analysis is conducted

**7) Number of Eligible Reporting Guidelines Used in Assessment of Included Studies** [textbox]

Note: Versions that differ by year count as the same guideline; extensions count as separate guidelines. All versions should be counted separately.

*Example: If a study includes PRISMA 2009 and PRISMA 2020, enter “1”*

*Example: If a study includes CONSORT 2010 and CONSORT Harms, enter “2”*

*Example: If a study only includes CONSORT Harm items, enter “1”*

**8) Name of Eligible Reporting Guidelines Used in Assessment of Included Studies** [textbox]

Note: Use acronyms and specify year and extension (if any)

**9) Name of Non-Eligible Reporting Guidelines Used in Assessment of Included Studies** [textbox]

Note: Use acronyms and specify year and extension (if any) – Enter “N/A” if not applicable

**10) Number of Publications Evaluated for Reporting** [textbox]

Note: Indicate the number of publications included for both eligible and ineligible reporting guidelines.

**11) Description of Main Eligibility Criteria of Included Studies** [checkbox]

Note: Can check more than one

- By reporting guideline (e.g., any study that mentions adhering to the PRISMA statement)
- By study design (e.g., RCTs, diagnostic test accuracy studies)
- By field of research (e.g., any study in rheumatology or dentistry)
- By patient population (e.g., people with scleroderma)
- By journal (e.g., all studies published in BMJ or a set of journals such as BMJ, JAMA, and NEJM)
- Other (Only include something that could potentially be used as a single eligibility criteria)
- By intervention type

**12) Rating Method Used** [dropdown]

Note: Do not consider "not applicable" as a rating option

- Dichotomous (e.g., yes/no)
- Multi-level (e.g., fully/partially/not reported)
- Other (Provide a brief description of rating method) [additional textbox]
- Not reported

**13) How many raters evaluated adherence to reporting guidelines?** [dropdown]

- 1 rater
- 1 rater with validation of all ratings from a second rater
- 1 rater with validation of some but not all ratings from a second rater
- 2 or more independent raters
  - Did they report concurrence between raters? If yes, provide a description of how agreement between raters was reported and result [additional textbox]
- 2 or more raters (independence not stated)
- Other [textbox]
- Not reported

**14) Did the authors provide an adequate coding explanation?** [dropdown/radio]

Coding Guide for Provision of an Adequate Coding Explanation

Fully Replicable: The authors provided a detailed description of what aspects needed to be present for each item and sub-item and explained how the presence or absence of these aspects led to the item rating (e.g., yes/no or fully/partially/not reported).

*Example^1-3^ using Item 6a of the CONSORT 2010 Checklis^4^: Completely defined pre-specified primary and secondary outcome measures, including how and when they were assessed*

*Fully: The authors clearly define the pre-specified primary and secondary outcome measures, including how and when they were assessed.*

*Partially: The authors only define the pre-specified primary and secondary outcome measures but not how and when they were assessed or they describe how and when outcomes were assessed but not the measures.*

*Not reported: The authors do not define the pre-specified primary and secondary outcome measures and do not define how and when they were assessed.*

Partially Replicable: The authors provided some detail regarding the aspects that needed to be present for each item and sub-item, but did not provide enough information to replicate.

Example

*Yes: The authors describe most components of each item needed to be present.*

*No: The authors do not describe the necessary components of each item.*

Not Replicable: The authors did not provide any explanation regarding how they rated reporting of each item.

- Fully replicable
- Partially replicable
- Not replicable

**15) Level of Reporting of Included Study Results** [dropdown/radio]

Note: If either of the “partially reported” options is chosen, provide details on level of reporting in the textbox, or reasoning behind why only some study outcomes were reported and others not.

*Example (Partially Reported – All Studies): Authors reported % CONSORT criteria of reporting quality fulfilled for each study.*

*Example (Partially Reported – Some Studies): Authors only reported complete outcomes for studies that adhered to at least half of reporting items.*

Coding Guide for Assessing Reporting of Included Study Results

Completely Reported: The authors provided ratings for each item from each individual study.

Partially Reported – All Studies: The authors provided some form of rating for each study but did not provide item-by-item results. This might include cases where the authors reported (1) a summary of the ratings for each individual study (e.g. 4/30 “yes” and 26/30 items “no") or (2) a summary for each individual study and item-by-item ratings for some items only.

Partially Reported – Some Studies: The authors reported ratings completely for some studies, but results for other studies were not reported or are partially reported.

Not Reported: The authors did not provide any results for individual studies.

- Completely reported
- Partially reported – All studies (Provide details on level of reporting) [additional textbox]
- Partially reported - Some studies (Provide explanation as to why some studies were reported and others not) [additional textbox]
- Not reported

**16) Publication Conclusion** [dropdown]

Coding Guide for Extracting Conclusions from Abstracts

Adequate: In the abstract conclusion, the authors state that overall adherence to reporting guidelines is at an adequate level.

Inadequate – Implicit: In the abstract conclusion, the authors state a recommendation that implies some degree of inadequacy, but there is no clear statement on the overall degree of adherence to reporting guidelines.

*Example:*

(1) *Completeness of reporting with respect to the PRISMA-DTA and PRISMA-DTA for Abstracts has improved modestly since the publication of the PRISMA-DTA guideline; however, increasing awareness of the specific weakness provides the chance for completeness improvement.*

Inadequate – Explicit: In the abstract conclusion, the authors state that overall adherence to reporting guidelines is inadequate.

*Examples:*

(1) *How administrative data are used in trials is often sub-optimally reported. CONSORT-ROUTINE uptake may improve reporting.*
(2) *Reporting of trials using registries was often poor, particularly details on data linkage and quality. Better reporting is needed for appropriate interpretation of the results of these trials.*
(3) *We found that the completeness of PRO reporting in RCTs involving AUD was deficient.*
(4) *The reporting quality of abstracts of RCTs presented at international cardiothoracic conferences is poor when benchmarked against the CONSORT-A standards.*

Mixed: In the abstract conclusion, the authors state that overall adherence to reporting guidelines is mixed.

*Examples:*

(1) *The reporting quality of SRs that underpin CPGs in breast cancer management widely varies.*
(2) *The quality of reporting of published massage RCTs is variable and in need of improvement.*

Vague: In the abstract conclusion, it is difficult to ascertain what the conclusion is regarding overall adherence to reporting guidelines. This might occur, for example, if the conclusion only comments on a small set of items.

*Example:*

(1) *Reporting quality of clinical studies had deficits in trial design-, recruitment-, allocation-, and outcome-related aspects.*

No Mention: In the abstract conclusion, there is no mention of adherence to reporting guidelines.

- Adequate
- Inadequate-Implicit
- Inadequate-Explicit
- Mixed
- Vague
- No mention

**17) Notes** [textbox]

**Supplementary Material 3: Coding Guide for Provision of an Adequate Coding Explanation**

**Fully Replicable:** The authors provided a detailed description of what aspects needed to be present for each item and sub-item and explained how the presence or absence of these aspects led to the item rating (e.g., yes/no or fully/partially/not reported).

Example^1-3^ using Item 6a of the CONSORT 2010 Checklist^4^: Completely defined pre-specified primary and secondary outcome measures, including how and when they were assessed

Fully: The authors clearly define the pre-specified primary and secondary outcome measures, including how and when they were assessed.

Partially: The authors only define the pre-specified primary and secondary outcome measures but not how and when they were assessed or they describe how and when outcomes were assessed but not the measures.

Not reported: The authors do not define the pre-specified primary and secondary outcome measures and do not define how and when they were assessed.

**Partially Replicable:** The authors provided some detail regarding the aspects that needed to be present for each item and sub-item, but did not provide enough information to replicate.

Example

Yes: The authors describe most components of each item needed to be present.

No: The authors do not describe the necessary components of each item.

**Not Replicable:** The authors did not provide any explanation regarding how they rated reporting of each item.

**REFERENCES: Supplementary Materials 2 and 3**

1. Imran M, Mc Cord KA, McCall SJ, et al. Reporting transparency and completeness in trials: paper 3 – trials conducted using administrative databases do not adequately report elements related to use of databases. *J Clin Epidemiol* 2022;141:187-197.
2. McCall SJ, Imran M, Hemkens LG, et al. Reporting transparency and completeness in trials: paper 4 – reporting of randomised controlled trials conducted using routinely collected electronic records – room for improvement. *J Clin Epidemiol* 2022;141:198-209.
3. McCord KA, Imran M, Rice DB, et al. Reporting transparency and completeness in trials: Paper 2 – reporting of randomised trials using registries was often inadequate and hindered the interpretation of results. *J Clin Epidemiol* 2022;141:175-186.
4. Schulz KF, Altman DG, Moher D, et al. CONSORT 2010 statement: updated guidelines for reporting parallel group randomised trials. *BMJ* 2010;340:c332.

**Supplementary Material 4: Coding Guide for Assessing Reporting of Included Study Results**

**Completely Reported:** The authors provided ratings for each item from each individual study.

**Partially Reported – All Studies:** The authors provided some form of rating for each study, but did not provide item-by-item results. This might include cases where the authors reported (1) a summary of the ratings for each individual study (e.g. 4/30 “yes” and 26/30 items “no") or (2) a summary for each individual study and item-by-item ratings for some items only.

**Partially Reported – Some Studies:** The authors reported ratings completely for some studies, but results for other studies were not reported or are partially reported.

**Not Reported:** The authors did not provide any results for individual studies.

**Supplementary Material 5: Characteristics of Included Studies**

|  |  |  |  |  |  |  |  |  | **Eligibility Criteria of Included Studies** | | | | | | |
| --- | --- | --- | --- | --- | --- | --- | --- | --- | --- | --- | --- | --- | --- | --- | --- |
| **First Author** | **Year**^a^ | **Journal** | **Journal Impact Factor** | **Country** | **Research Question** | **Number of Eligible Reporting Guidelines** | **Name of Eligible Reporting Guidelines** | **Number of Publications** | **Study Design** | **Field of Research** | **Patient Population** | **Journal** | **Intervention Type** | **Included in specified guidelines** | **Other** |
| Alharbi^S1^ | 2020 | Contemporary Clinical Trials Communications | No impact factor | Saudi Arabia | Reporting only | 1 | CONSORT-A | 177 | ✓ |  |  | ✓ |  |  |  |
| Candela^S2^ | 2020 | International Journal of Environmental Research and Public Health | 4.6 | Italy | Main reporting | 1 | CONSORT | 183 | ✓ |  | ✓ |  |  |  |  |
| Dai^S3^ | 2020 | BMC Medical Research Methodology | 4.6 | China | Reporting only | 1 | STROBE | 165 | ✓ |  |  |  |  |  |  |
| Duan^S4^ | 2020 | BMC Complementary Medicine and Therapies | 2.8 | China | Main unclear | 1 | STROBE | 199 | ✓ |  |  |  | ✓ |  |  |
| Gore^S5^ | 2020 | Journal of Chronic Obstructive Pulmonary Disease | 2.1 | United States | Main not reporting | 1 | STROBE | 12 | ✓ |  | ✓ |  |  |  |  |
| Gundogan^S6^ | 2020 | JAAD International | No impact factor | United Kingdom | Reporting only | 1 | PRISMA | 166 | ✓ | ✓ |  | ✓ |  |  |  |
| Hogan^S7^ | 2020 | American Journal of Clinical Pathology | 5.4 | United States | Reporting only | 1 | STARD | 171 | ✓ | ✓ |  |  |  |  |  |
| Hou^S8^ | 2020 | Evidence-Based Complementary and Alternative Medicine | 2.7 | China | Main unclear | 1 | PRISMA | 11 | ✓ |  | ✓ |  | ✓ |  |  |
| Huang^S9^ | 2020 | Frontiers in Aging Neuroscience | 5.7 | China | Main unclear | 1 | PRISMA | 11 | ✓ |  | ✓ |  | ✓ |  |  |
| Huang^S10^ | 2020 | Journal of Pain Research | 2.8 | China | Main unclear | 1 | PRISMA | 19 | ✓ |  | ✓ |  | ✓ |  |  |
| Li^S11^ | 2020 | Evidence-Based Complementary and Alternative Medicine | 2.7 | China | Reporting only | 2 | CONSORT-CHM | 144 | ✓ |  | ✓ |  | ✓ |  |  |
| Li^S12^ | 2020 | Journal of Dentistry | 5.0 | China | Reporting only | 1 | PRISMA-A | 160 | ✓ |  |  |  | ✓ |  |  |
| Lyu^S13^ | 2020 | Gastroenterology Research and Practice | 1.9 | China | Main unclear | 1 | PRISMA | 33 | ✓ |  | ✓ |  |  |  |  |
| Manouchehri^S14^ | 2020 | Iranian Journal of Nursing and Midwifery Research | No impact factor | Iran | Reporting only | 1 | CONSORT | 30 | ✓ |  | ✓ |  | ✓ |  |  |
| Mazhar^S15^ | 2020 | International Journal of Infectious Diseases | 12.1 | United Kingdom | Main unclear | 1 | CONSORT-Harms (Extension items only) | 16 | ✓ |  | ✓ |  | ✓ |  |  |
| Ngah^S16^ | 2020 | Vaccines (Basel) | 5.0 | South Africa | Reporting only | 1 | CONSORT | 124 |  |  |  |  | ✓ |  |  |
| Rainkie^S17^ | 2020 | PLoS One | 3.8 | Qatar | Main unclear | 2 | PRISMA and PRISMA-P | 51 | ✓ |  | ✓ |  |  |  |  |
| Suchá^S18^ | 2020 | Radiology: Cardiothoracic Imaging | No impact factor | the Netherlands | Main unclear | 1 | STARD | 13 | ✓ |  | ✓ |  |  |  |  |
| Tian^S19^ | 2020 | Chinese Medicine | 4.5 | China | Main unclear | 1 | PRISMA | 97 | ✓ |  |  |  | ✓ |  |  |
| Zhang^S20^ | 2020 | Chinese Acupuncture & Moxibustion | No impact factor | China | Reporting only | 2 | CONSORT  and  STRICTA | 33 | ✓ |  | ✓ |  | ✓ |  |  |
| Zhu^S21^ | 2020 | International Immunopharmacology | 5.7 | China | Main unclear | 1 | PRISMA | 11 | ✓ |  | ✓ |  | ✓ |  |  |
| Adams^S22^ | 2021 | BMJ Open | 3.0 | United States | Main unclear | 3 | CONSORT, CONSORT-NPT, CONSORT-Harms,  and  TIDieR | 96 | ✓ |  | ✓ |  | ✓ |  |  |
| Adobes Martin^S23^ | 2021 | BMC Medical Research Methodology | 4.6 | Spain | Reporting only | 1 | PRISMA-A | 265 | ✓ | ✓ |  |  |  |  |  |
| Almeheyawi^S24^ | 2021 | Journal of Foot and Ankle Research | 3.1 | Saudi Arabia | Main not reporting | 1 | STROBE | 39 | ✓ |  | ✓ |  |  |  |  |
| Bachelet^S25^ | 2021 | BMC Medical Research Methodology | 4.6 | Chile | Main unclear | 1 | CONSORT | 392 | ✓ | ✓ |  | ✓ |  |  |  |
| Bae^S26^ | 2021 | Complementary Therapies in Clinical Practice | 3.6 | South Korea | Reporting only | 2 | PRISMA and PRISMA-NMA | 42 | ✓ |  |  |  | ✓ |  |  |
| Barhli^S27^ | 2021 | Critical Reviews in Oncology / Hematology | 6.6 | France | Reporting only | 1 | CONSORT-Harms (Extension items only) | 46 | ✓ |  | ✓ |  | ✓ |  |  |
| Beneki^S28^ | 2021 | Journal of Thrombosis and Thrombolysis | 5.2 | Greece | Reporting only | 1 | CONSORT | 13 | ✓ |  | ✓ |  | ✓ |  |  |
| Berton^S29^ | 2021 | Journal of Clinical Medicine | 5.0 | Italy | Reporting only | 1 | CONSORT | 79 | ✓ |  | ✓ |  |  |  |  |
| Besag^S30^ | 2021 | Journal of Psychopharmacology | 4.6 | United Kingdom | Main not reporting | 1 | CONSORT | 17 | ✓ |  | ✓ |  | ✓ |  |  |
| Bruno^S31^ | 2021 | American Journal of Obstetrics & Gynecology MFM | 8.7 | United States | Main unclear | 1 | CONSORT | 170 | ✓ |  |  | ✓ |  |  |  |
| Burton^S32^ | 2021 | Cochrane Database of Systematic Reviews | 12.0 | United Kingdom | Main not reporting | 1 | STARDdem | 13 | ✓ |  | ✓ |  |  |  | IQCODE had to be used as an informant questionnaire |
| Canagarajah^S33^ | 2021 | European Archives of Oto-Rhino-Laryngology | 3.2 | United Kingdom | Reporting only | 1 | CONSORT | 41 | ✓ |  |  |  | ✓ |  |  |
| Cao^S34^ | 2021 | Journal of Clinical Epidemiology | 7.4 | China | Main unclear | 1 | PRISMA | 336 | ✓ |  |  |  |  |  |  |
| Chair^S35^ | 2021 | International Journal of Environmental Research and Public Health | 4.6 | China | Main not reporting | 1 | STROBE | 61 | ✓ |  | ✓ |  |  |  |  |
| Cho^S36^ | 2021 | Healthcare (Basel) | 3.2 | Korea | Reporting only | 1 | PRISMA | 47 | ✓ |  |  | ✓ |  |  |  |
| Du^S37^ | 2021 | Thoracic Cancer | 3.2 | China | Reporting only | 1 | CONSORT^c^ | 152 | ✓ |  | ✓ |  | ✓ |  |  |
| Duan^S38^ | 2021 | The American Journal of Chinese Medicine | 6.0 | China | Main unclear | 1 | STROBE (Cross-sectional studies) | 198 |  | ✓ |  |  |  |  |  |
| Eliya^S39^ | 2021 | Journal of the American Heart Association | 6.1 | Canada | Main unclear | 1 | CONSORT PRO | 226 | ✓ |  | ✓ |  |  |  |  |
| Escobar-Viera^S40^ | 2021 | Internet Interventions | 5.4 | United States | Main unclear | 2 | CONSORT and CONSORT for pilot and feasibility trials | 18 |  |  | ✓ |  | ✓ |  |  |
| Garnier^S41^ | 2021 | Neuro-Oncology Practice | No impact factor | France | Reporting only | 1 | CONSORT-PRO (Extension items only) | 43 | ✓ |  | ✓ |  |  |  |  |
| Hu^S42^ | 2021 | Journal of Advanced Nursing | 3.1 | China | Main not reporting | 1 | PRISMA | 10 | ✓ |  |  |  | ✓ |  |  |
| Huang^S43^ | 2021 | Pain Research and Management | 2.7 | China | Main unclear | 1 | PRISMA | 10 | ✓ |  | ✓ |  | ✓ |  |  |
| Jacobsen^S44^ | 2021 | British Journal of Anaesthesia | 11.7 | United States | Main unclear | 1 | PRISMA | 78 | ✓ |  |  |  |  | ✓ |  |
| Jamnani^S45^ | 2021 | Medical Journal of the Islamic Republic of Iran | No impact factor | Iran | Main not reporting | 1 | STROBE | 28 |  |  |  |  | ✓ |  | Studies of willingness to pay and acceptability of cervical cancer prevention |
| Jumah^S46^ | 2021 | Stroke | 10.2 | United States | Reporting only | 2 | PRISMA-IPD | 31 | ✓ |  | ✓ |  |  |  |  |
| Kennedy^S47^ | 2021 | Haemophilia | 4.3 | Ireland | Main not reporting | 1 | STROBE | 36 | ✓ |  | ✓ |  |  |  |  |
| Kim^S48^ | 2021 | Harm Reduction Journal | 4.8 | United States | Main unclear | 1 | PRISMA | 20 | ✓ |  |  |  | ✓ |  |  |
| Knippschild^S49^ | 2021 | BMJ Open | 3.0 | Germany | Reporting only | 1 | CONSORT-A | 212 | ✓ | ✓ |  |  |  |  |  |
| Kshirsagar^S50^ | 2021 | Journal of Oral and Maxillofacial Surgery | 2.1 | India | Reporting only | 2 | CONSORT and CONSORT Abstracts | 80 | ✓ |  | ✓ |  |  |  |  |
| Li^S51^ | 2021 | Annals of Palliative Medicine | 1.9 | China | Main unclear | 1 | PRISMA | 12 | ✓ |  | ✓ |  | ✓ |  |  |
| Li^S52^ | 2021 | Evidence-Based Complementary and Alternative Medicine | 2.7 | China | Main unclear | 1 | CONSORT | 39 | ✓ |  |  |  | ✓ |  |  |
| Li^S53^ | 2021 | Expert Review of Molecular Diagnostics | 5.7 | China | Main not reporting | 2 | PRISMA-DTA | 14 | ✓ |  | ✓ |  |  |  |  |
| Li^S54^ | 2021 | Journal of Clinical Epidemiology | 7.4 | China | Main unclear | 1 | PRISMA^c^ | 243 | ✓ |  |  |  |  |  |  |
| Liang^S55^ | 2021 | Chinese Medicine | 4.5 | Macao | Main unclear | 2 | CONSORT-CHM | 53 | ✓ |  | ✓ |  | ✓ |  |  |
| Liu^S56^ | 2021 | Journal of Pain Research | 2.8 | China | Reporting only | 2 | CONSORT and STRICTA | 31 | ✓ |  |  |  | ✓ |  |  |
| Liu^S57^ | 2021 | Obesity Reviews | 10.9 | China | Reporting only | 2 | CONSORT and CONSORT-NPT | 102 | ✓ |  |  |  | ✓ |  |  |
| Lu^S58^ | 2021 | Phytomedicine | 6.7 | China | Main unclear | 1 | PRISMA | 19 | ✓ |  |  |  | ✓ |  |  |
| Malone^S59^ | 2021 | Cancer Medicine | 4.7 | Canada | Reporting only | 2 | CONSORT and CONSORT-PRO | 33 | ✓ |  |  |  | ✓ |  |  |
| McGrath^S60^ | 2021 | Journal of Pediatric Urology | 1.9 | Canada | Reporting only | 1 | CONSORT extension checklist for pilot studies | 36 | ✓ | ✓ | ✓ |  |  |  |  |
| Morand^S61^ | 2021 | Archives de Pédiatrie | 1.8 | France | Main unclear | 1 | STROBE^c^ | 52 |  |  | ✓ |  |  |  |  |
| Nascimento^S62^ | 2021 | Brazilian Journal of Physical Therapy | 4.8 | Brazil | Main unclear | 1 | PRISMA-A | 66 | ✓ |  |  |  | ✓ |  |  |
| Prager^S63^ | 2021 | BMJ Evidence-Based Medicine | 4.7 | Canada | Reporting only | 3 | PRISMA-DTA  and PRISMA-DTA for Abstracts | 71 | ✓ |  |  |  |  |  |  |
| Prins^S64^ | 2021 | Archives of Disease in Childhood | 5.0 | the Netherlands | Reporting only | 1 | CONSORT-Harms (Extensions items only) | 100 | ✓ |  | ✓ |  | ✓ |  |  |
| Qin^S65^ | 2021 | European Journal of Orthodontics | 3.1 | China | Reporting only | 2 | CONSORT and CONSORT for within person randomised trials | 42 | ✓ |  |  | ✓ | ✓ |  |  |
| Rod^S66^ | 2021 | Accident Analysis and Prevention | 6.4 | Australia | Main not reporting | 1 | STROBE | 60 |  |  | ✓ |  |  |  |  |
| Shi^S67^ | 2021 | Systematic Reviews | 3.1 | China | Main not reporting | 1 | PRISMA | 31 | ✓ |  | ✓ |  |  |  |  |
| Sun^S68^ | 2021 | Nursing Open | 1.9 | China | Main unclear | 1 | PRISMA | 130 | ✓ |  | ✓ |  | ✓ |  |  |
| Tian^S69^ | 2021 | Evidence-Based Complementary and Alternative Medicine | 2.7 | China | Main unclear | 1 | PRISMA | 14 | ✓ |  | ✓ |  | ✓ |  |  |
| Veroniki^S70^ | 2021 | Systematic Reviews | 3.1 | Greece | Main reporting | 2 | PRISMA-NMA | 1,144 | ✓ |  |  |  | ✓ |  |  |
| Wenhui^S71^ | 2021 | Diabetes Research and Clinical Practice | 8.2 | China | Main not reporting | 1 | PRISMA | 11 | ✓ |  | ✓ |  | ✓ |  |  |
| Wright^S72^ | 2021 | Clinical Medicine & Research | No impact factor | United States | Main reporting | 1 | STARD | 26 | ✓ |  |  |  |  | ✓ |  |
| Yang^S73^ | 2021 | International Journal of General Medicine | 2.1 | China | Main unclear | 1 | PRISMA | 12 | ✓ |  | ✓ |  | ✓ |  |  |
| Yin^S74^ | 2021 | PLoS One | 3.8 | China | Reporting only | 1 | CONSORT | 53 | ✓ |  | ✓ |  |  |  |  |
| Yuan^S75^ | 2021 | Evidence-Based Complementary and Alternative Medicine | 2.7 | China | Main unclear | 2 | PRISMA-NMA | 29 | ✓ |  |  |  | ✓ |  |  |
| Zhang^S76^ | 2021 | Asian Pacific Journal of Clinical Nutrition | No impact factor | China | Reporting only | 2 | STROBE-nut | 200 | ✓ |  |  | ✓ |  |  |  |
| Zhang^S77^ | 2021 | Chinese Medicine | 4.5 | China | Main unclear | 2 | CONSORT and CONSORT non-pharmacological treatment interventions | 2,447 | ✓ |  |  |  | ✓ |  |  |
| Zhang^S78^ | 2021 | Frontiers in Pharmacology | 6.0 | China | Main unclear | 1 | PRISMA | 14 | ✓ |  | ✓ |  | ✓ |  |  |
| Zhang^S79^ | 2021 | Journal of Clinical Epidemiology | 7.4 | China^b^ | Main unclear | 1 | CONSORT | 2,844 | ✓ |  |  |  |  |  |  |
| Zheng^S80^ | 2021 | Annals of Translational Medicine | 3.6 | China | Reporting only | 1 | STARD | 45 | ✓ |  |  | ✓ |  |  |  |
| Zheng^S81^ | 2021 | Frontiers in Medicine | 5.1 | China | Main unclear | 1 | PRISMA | 238 | ✓ |  |  |  | ✓ |  |  |
| Al-Abedalla^S82^ | 2022 | JDR Clinical & Translational Research | No impact factor | United States | Main unclear | 1 | CONSORT | 32 | ✓ |  |  |  | ✓ |  |  |
| Alshahwani^S83^ | 2022 | Journal of Surgical Research | 2.4 | Qatar | Main  unclear | 1 | PRISMA | 21 | ✓ |  | ✓ |  |  |  |  |
| Astur^S84^ | 2022 | einstein | No impact factor | Brazil | Main reporting | 1 | PRISMA | 65 | ✓ |  |  |  | ✓ |  |  |
| Batioja^S85^ | 2022 | Journal of Pediatric Orthopaedics | 2.5 | United States | Main unclear | 1 | CONSORT | 23 | ✓ |  |  |  |  | ✓ |  |
| Bole^S86^ | 2022 | The Journal of Sexual Medicine | 3.9 | United States | Main unclear | 1 | PRISMA | 18 | ✓ |  | ✓ |  |  |  |  |
| Bonetti^S87^ | 2022 | Research in Social and Administrative Pharmacy | 3.3 | Portugal | Main unclear | 1 | PRISMA | 109 | ✓ |  |  |  | ✓ |  |  |
| Cervantes^S88^ | 2022 | Psychiatric Services | 4.2 | United States | Main not reporting | 1 | STROBE | 11 |  |  | ✓ |  |  |  |  |
| Chen^S89^ | 2022 | Journal of Ginseng Research | 5.7 | Macao | Main unclear | 1 | CONSORT | 91 | ✓ |  |  |  | ✓ |  |  |
| Cindro^S90^ | 2022 | BMJ Open | 3.0 | Croatia | Main reporting | 1 | CONSORT-A | 451 | ✓ |  | ✓ |  |  |  |  |
| de Lucena Alves^S91^ | 2022 | Clinical Implant Dentistry and Related Research | 4.3 | Brazil | Main unclear | 1 | PRISMA-Abstracts^c^ | 45 | ✓ |  | ✓ |  | ✓ |  |  |
| Douglas^S92^ | 2022 | BMJ Evidence-Based Medicine | 4.7 | United States | Main reporting | 1 | CONSORT-PRO | 19 | ✓ |  | ✓ |  |  |  |  |
| Feng^S93^ | 2022 | Clinical and Translational Gastroenterology | 4.4 | the Netherlands | Main not reporting | 1 | STARD | 136 |  |  | ✓ |  |  |  |  |
| Fernández-Pires^S94^ | 2022 | The American Journal of Occupational Therapy | 2.8 | Spain | Reporting only | 1 | CONSORT-A | 78 | ✓ |  |  | ✓ |  |  |  |
| Frank^S95^ | 2022 | Journal of Magnetic Resonance Imaging | 5.1 | Canada | Main reporting | 1 | STARD for Abstracts | 2000 | ✓ |  |  | ✓ |  |  | Abstracts submitted to specified conferences |
| Gebran^S96^ | 2022 | Journal of the American College of Surgeons | 6.5 | United States | Reporting only | 1 | RECORD | 118 |  |  |  |  |  |  | Publications from a specified program |
| Gupta^S97^ | 2022 | Perspectives in Clinical Research | No impact factor | India | Main reporting | 1 | CONSORT | 28 | ✓ |  |  | ✓ |  |  |  |
| Helliwell^S98^ | 2022 | Health Science Reports | No impact factor | United Kingdom | Main unclear | 1 | PRISMA | 31 | ✓ |  |  |  | ✓ |  |  |
| Huang^S99^ | 2022 | Frontiers in Public Health | 6.5 | China | Main unclear | 1 | PRISMA | 10 | ✓ |  | ✓ |  | ✓ |  |  |
| Imran^S100^ | 2022 | Journal of Clinical Epidemiology | 7.4 | Canada | Main unclear | 1 | CONSORT-ROUTINE | 33 | ✓ |  |  |  |  |  |  |
| Jung^S101^ | 2022 | Clinical Microbiology and Infection | 13.3 | South Korea | Reporting only | 1 | CONSORT | 87 | ✓ |  | ✓ |  | ✓ |  |  |
| Kanukula^S102^ | 2022 | BMJ Evidence-Based Medicine | 4.7 | Australia | Main unclear | 1 | PRISMA | 40 | ✓ |  |  |  |  | ✓ |  |
| Kazi^S103^ | 2022 | Journal of Magnetic Resonance Imaging | 5.1 | Canada | Main unclear | 2 | STARD  and  STARD for Abstracts | 84 | ✓ |  |  | ✓ |  |  |  |
| Khachfe^S104^ | 2022 | The American Journal of Surgery | 3.1 | United States | Main unclear | 2 | STROBE and RECORD | 86 |  |  | ✓ |  |  |  | Publications using specified database |
| Khan^S105^ | 2022 | Multiple Sclerosis and Related Disorders | 4.8 | United States | Reporting only | 1 | CONSORT-PRO | 92 | ✓ |  | ✓ |  |  |  |  |
| Kim^S106^ | 2022 | Frontiers in Medicine | 5.1 | South Korea | Main not reporting | 1 | STROBE | 62 | ✓ |  |  |  | ✓ |  |  |
| Labiste^S107^ | 2022 | Skeletal Radiology | 2.1 | United States | Main not reporting | 1 | STARD | 21 | ✓ |  | ✓ |  |  |  |  |
| Li^S108^ | 2022 | Evidence-Based Complementary and Alternative Medicine | 2.7 | China | Main not reporting | 1 | PRISMA | 27 | ✓ |  | ✓ |  |  |  |  |
| Love^S109^ | 2022 | Critical Reviews in Oncology / Hematology | 6.6 | United States | Reporting only | 1 | PRISMA | 109 | ✓ |  |  |  |  | ✓ |  |
| Lu^S110^ | 2022 | Journal of Integrative Medicine | 4.0 | China | Main unclear | 1 | PRISMA | 20 | ✓ |  |  |  | ✓ |  |  |
| McCall^S111^ | 2022 | Journal of Clinical Epidemiology | 7.4 | United Kingdom | Main unclear | 2 | CONSORT and CONSORT-ROUTINE | 60 | ✓ |  |  |  |  |  |  |
| Mc Cord^S112^ | 2022 | Journal of Clinical Epidemiology | 7.4 | Switzerland | Main unclear | 1 | CONSORT-ROUTINE | 47 | ✓ |  |  |  |  |  |  |
| McErlean^S113^ | 2022 | Irish Journal of Medical Science | 2.1 | United Kingdom | Reporting only | 1 | CONSORT | 50 | ✓ |  |  | ✓ |  |  |  |
| Menne^S114^ | 2022 | Journal of Periodontology | 4.5 | Germany | Reporting only | 1 | CONSORT-A | 434 | ✓ | ✓ |  | ✓ |  |  |  |
| Newman^S115^ | 2022 | Diabetes Research and Clinical Practice | 8.2 | Ireland | Main not reporting | 2 | CONSORT and CONSORT-PRO | 206 | ✓ |  | ✓ |  |  |  |  |
| Park^S116^ | 2022 | Korean Journal of Radiology | 7.1 | Korea | Reporting only | 2 | PRISMA and PRISMA-A | 24 | ✓ |  |  | ✓ |  |  |  |
| Peña^S117^ | 2022 | Urology | 2.6 | United States | Main unclear | 1 | PRISMA | 120 | ✓ | ✓ |  |  |  | ✓ |  |
| Pfannenstiel^S118^ | 2022 | The International Journal of Oral & Maxillofacial Implants | 2.9 | Germany | Main reporting | 2 | CONSORT for within person randomised trials | 244 | ✓ |  |  |  | ✓ |  |  |
| Ruan^S119^ | 2022 | International Journal of General Medicine | 2.1 | China | Reporting only | 2 | STRICTA | 44 | ✓ |  | ✓ |  | ✓ |  |  |
| Shi^S120^ | 2022 | Cardiology Research and Practice | 2.0 | China | Main unclear | 1 | PRISMA | 12 | ✓ |  | ✓ |  | ✓ |  |  |
| Shi^S121^ | 2022 | Drug Design, Development and Therapy | 4.3 | China | Main not reporting | 1 | PRISMA | 13 | ✓ |  | ✓ |  | ✓ |  |  |
| Shin^S122^ | 2022 | International Journal of Environmental Research and Public Health | 4.6 | Korea | Main unclear | 1 | PRISMA | 41 | ✓ | ✓ |  |  |  |  |  |
| Siew^S123^ | 2022 | Psychoneuroendocrinology | 4.7 | Australia | Main not reporting | 1 | STROBE | 10 | ✓ |  |  |  | ✓ |  |  |
| Streck^S124^ | 2022 | Nicotine and Tobacco Research | 5.8 | United States | Main unclear | 1 | PRISMA | 98 | ✓ |  |  |  |  | ✓ |  |
| Stunnenberg^S125^ | 2022 | Neurology | 11.8 | the Netherlands | Main unclear | 2 | CONSORT and  CENT | 40 | ✓ | ✓ |  |  |  |  |  |
| Tanner^S126^ | 2022 | Drug and Alcohol Dependence | 4.9 | United States | Main reporting | 1 | PRISMA | 98 | ✓ |  |  |  |  | ✓ |  |
| Torgerson^S127^ | 2022 | International Journal of Pediatric Otorhinolaryngology | 1.6 | United States | Main unclear | 1 | PRISMA | 80 | ✓ |  |  |  |  | ✓ |  |
| Warrier^S128^ | 2022 | Perspectives in Clinical Research | No impact factor | India | Main reporting | 1 | CONSORT | 276 | ✓ |  |  | ✓ |  |  |  |
| Yang^S129^ | 2022 | Evidence-Based Complementary and Alternative Medicine | 2.7 | China | Reporting only | 2 | CONSORT and STRICTA | 102 | ✓ |  |  |  | ✓ |  |  |
| Yang^S130^ | 2022 | Journal of Ethnopharmacology | 5.2 | China | Main unclear | 1 | PRISMA | 52 | ✓ |  | ✓ |  | ✓ |  |  |
| Yao^S131^ | 2022 | Journal of Integrative Medicine | 4.0 | China | Main unclear | 1 | PRISMA-A | 13 | ✓ |  | ✓ |  | ✓ |  |  |
| Yin^S132^ | 2022 | Frontiers in Physiology | 4.8 | China | Main not reporting | 1 | PRISMA | 10 | ✓ |  | ✓ |  | ✓ |  |  |
| Yin^S133^ | 2022 | International Journal of Infectious Diseases | 12.1 | China | Main reporting | 1 | CONSORT-A | 53 | ✓ |  | ✓ |  |  |  |  |
| Yuniar^S134^ | 2022 | Vaccines (Basel) | 5.0 | Korea | Main reporting | 1 | CONSORT Harms | 61 | ✓ |  |  |  | ✓ |  |  |
| Zhang^S135^ | 2022 | Human Vaccines & Immunotherapeutics | 4.5 | China | Reporting only | 2 | CONSORT and CONSORT Harms | 22 | ✓ |  |  |  | ✓ |  |  |
| Zhou^S136^ | 2022 | Therapeutic Advances in Gastroenterology | 4.8 | China | Main unclear | 3 | CONSORT extension to randomised crossover trials  and CONSORT-A | 173 | ✓ |  |  | ✓ |  |  |  |
| Ziemann^S137^ | 2022 | BMC Medical Research Methodology | 4.6 | Germany | Reporting only | 1 | STROBE | 147 | ✓ |  | ✓ |  |  |  |  |
| Anderson^S138^ | Online only | The Journal of Arthroplasty | 4.4 | United States | Reporting only | 1 | CONSORT Harms | 173 | ✓ |  |  |  |  | ✓ |  |
| Bonafiglia^S139^ | Online only | Journal of Sport and Health Science | 13.1 | Canada | Main not reporting | 2 | CONSORT Extension for Nonpharmacologic Trial Abstracts | 27 |  |  |  |  | ✓ |  |  |
| Brito-Suárez^S140^ | Online only | Pediatric Hematology and Oncology | 2.1 | Mexico | Main not reporting | 1 | STROBE | 10 | ✓ |  | ✓ |  |  |  |  |
| Cremades-Martínez^S141^ | Online only | Microbiology Spectrum | 9.0 | Spain | Main unclear | 1 | STARD | 23 | ✓ |  | ✓ |  |  |  |  |
| Dhillon^S142^ | Online only | The Laryngoscope | 3.0 | United States | Main unclear | 1 | PRISMA | 142 | ✓ |  |  |  |  | ✓ |  |
| Garrett^S143^ | Online only | BMJ Evidence-Based Medicine | 4.7 | United States | Main unclear | 1 | PRISMA | 55 | ✓ |  |  |  |  | ✓ |  |
| Gysling^S144^ | Online only | Seminars in Thoracic and Cardiovascular Surgery | 2.4 | United Kingdom | Reporting only | 1 | CONSORT-A | 100 | ✓ |  |  |  |  |  | Abstracts presented at specified annual meetings |
| Innocenti^S145^ | Online only | Archives of Physical Medicine and Rehabilitation | 4.1 | the Netherlands | Main reporting | 1 | CONSORT | 200 | ✓ | ✓ |  |  |  |  |  |
| Kim^S146^ | Online only | Academic Radiology | 5.5 | South Korea | Reporting only | 2 | PRISMA-DTA  and PRISMA-DTA for Abstracts | 183 | ✓ | ✓ |  |  |  |  |  |
| Snider^S147^ | Online only | Clinical Breast Cancer | 3.1 | United States | Main unclear | 1 | PRISMA | 59 | ✓ |  |  |  |  | ✓ |  |
| Tosatto^S148^ | Online only | Restorative Neurology and Neuroscience | 3.0 | Italy | Main unclear | 1 | CONSORT-A | 120 | ✓ |  | ✓ |  |  |  |  |

^a^Year of publication based on when data were extracted

^b^This study has two corresponding authors; we selected the country of the corresponding author who was also the last author (Bian)

^c^Included additional reporting guidelines not eligible for our review.

**Supplementary Material 6: Rating Method and Number of Raters for Overall Sample and Subgroups**

|  | **N**  **% (95%CI)** | | | | | | | | | |
| --- | --- | --- | --- | --- | --- | --- | --- | --- | --- | --- |
| **Subgroups** | **Rating Method** | | | | **Number of Raters** | | | | | |
|  | Dichotomous | Multi-level | Other | Not  reported | 1 rater | 1 rater with validation from a second rater | 2 or more independent raters | 2 or more raters (independence not stated) | Other | Not Reported |
| **All** | 66  45% (37%, 53%) | 61  41% (34%, 49%) | 2  1% (0%, 5%) | 19  13% (8%, 19%) | 3  2% (1%, 6%) | 10  7% (4%, 12%) | 113  76% (69%, 83%) | 9  6% (3%, 11%) | 3  2% (1%, 6%) | 10  7% (4%, 12%) |
| **Country** |  |  |  |  |  |  |  |  |  |  |
| Canada | 6  75% (41%, 93%) | 1  13% (2%, 47%) | 0  0% (0%, 32%) | 1  13% (2%, 47%) | 0  0% (0%, 32%) | 1  13% (2%, 47%) | 6  75% (41%, 93%) | 0  0% (0%, 32%) | 0  0% (0%, 32%) | 1  13% (2%, 47%) |
| China | 14  28% (17%, 41%) | 33  65% (51%, 76%) | 0  0% (0%, 7%) | 4  8% (3%, 19%) | 0  0% (0%, 7%) | 2  4% (1%, 13%) | 46  90% (79%, 96%) | 3  6% (2%, 16%) | 0  0% (0%, 7%) | 0  0% (0%, 7%) |
| United Kingdom | 3  33% (12%, 65%) | 4  44% (19%, 73%) | 0  0% (0%, 30%) | 2  22% (6%, 55%) | 0  0% (0%, 30%) | 3  33% (12%, 65%) | 5  56% (27%, 81%) | 0  0% (0%, 30%) | 0  0% (0%, 30%) | 1  11% (2%, 44%) |
| United States | 10  37% (22%, 56%) | 12  44% (28%, 63%) | 1  4% (1%, 18%) | 4  15 (6%, 33%) | 1  4% (1%, 18%) | 1  4% (1%, 18%) | 21  78% (59%, 89%) | 4  15% (6%, 33%) | 0  0% (0%, 13%) | 0  0% (0%, 13%) |
| Other | 33  62% (49%, 74%) | 11  21% (12%, 34%) | 1  2% (0%, 10%) | 8  15% (8%, 27%) | 2  4% (1%, 13%) | 3  6% (2%, 15%) | 35  66% (53%, 77%) | 2  4% (1%, 13%) | 3  6% (2%, 15%) | 8  15% (8%, 27%) |
| **Journal Impact Factor**^a^ |  |  |  |  |  |  |  |  |  |  |
| Impact factor ≤ 2.9 | 16  36% (23%, 50%) | 21  47% (33%, 61%) | 0  0% (0%, 8%) | 8  18% (9%, 31%) | 0  0% (0%, 8%) | 0  0% (0%, 8%) | 36  80 (66%, 89%) | 2  4% (1%, 15%) | 2  4% (1%, 15%) | 5  11% (5%, 24%) |
| Impact factor > 2.9 | 50  49% (39%, 58%) | 40  39% (30%, 49%) | 2  2% (1%, 7%) | 11  11% (6%, 18%) | 3  3% (1%, 8%) | 10  10% (5%, 17%) | 77  75% (66, 82%) | 7  7% (3%, 13%) | 1  1% (0%, 5%) | 5  5% (2%, 11%) |
| **Reporting Guideline** |  |  |  |  |  |  |  |  |  |  |
| CONSORT & extensions | 35  57% (45%, 69%) | 14  23% (14%, 35%) | 2  3% (1%, 11%) | 10  16% (9%, 28%) | 0  0% (0%, 6%) | 6  10% (5%, 20%) | 41  67% (55%, 78%) | 4  7% (3%, 16%) | 3  5% (2%, 14%) | 7  11% (6%, 22%) |
| PRISMA & extensions | 14  24% (15%, 36%) | 40  68% (55%, 78%) | 0  0% (0%, 6%) | 5  9% (4%, 18%) | 0  0% (0%, 6%) | 0  0% (0%, 6%) | 53  90% (80%, 95%) | 4  7% (3%, 16%) | 0  0% (0%, 6%) | 2  3% (1%, 12%) |
| STARD & extensions | 6  60% (31%, 83%) | 2  20% (6%, 51%) | 0  0% (0%, 28%) | 2  20% (6%, 51%) | 0  0% (0%, 28%) | 0  0% (0%, 28%) | 10  100% (72%, 100%) | 0  0% (0%, 28%) | 0  0% ()%, 28%) | 0  0% (0%, 28%) |
| STROBE & extensions | 11  61% (39%, 80%) | 5  28% (13%, 51%) | 0  0% (0%, 18%) | 2  11% (3%, 33%) | 3  17% (6%, 39%) | 4  22% (%, 45%) | 9  50% (29%, 71%) | 1  6% (1%, 26%) | 0  0% (0%, 18%) | 1  6% (1%, 26%) |
| **Research Question** |  |  |  |  |  |  |  |  |  |  |
| Reporting is the only research question or there are multiple research questions and the main one is reporting or not defined | 55  44% (36%, 53%) | 50  40% (32%, 49%) | 2  2% (0%, 6%) | 17  14% (9%, 21%) | 0  0% (0%, 3%) | 7  6% (3%, 11%) | 96  77% (69%, 84%) | 9  7% (4%, 13%) | 3  2% (1%, 7%) | 9  7% (4%, 13%) |
| Reporting is not the main research question | 11  46% (28%, 65%) | 11  46% (28%, 65%) | 0  0% (0%, 14%) | 2  8% (2%, 26%) | 3  13% (4%, 31%) | 3  13% (4%, 31%) | 17  71% (51%, 85%) | 0  0% (0%, 14%) | 0  0% (0%, 14%) | 1  4% (1%, 20%) |

^a^Journals without an impact factor coded as 0.

**Supplementary Material 7: Outcomes for All Included Studies**

| **First Author** | **Year**^a^ | **Journal** | **Rating Method Used** | **Number of Raters** | **Concurrence Between Raters** | **Coding Explanation** | **Level of Reporting of Included Study Results** | **Publication Conclusion** |
| --- | --- | --- | --- | --- | --- | --- | --- | --- |
| Alharbi^S1^ | 2020 | Contemporary Clinical Trials Communications | Dichotomous | Other (A random sample of 10% of the papers was scored by a second examiner) | N/A | Not replicable | Not reported | Inadequate - Explicit |
| Candela^S2^ | 2020 | International Journal of Environmental Research and Public Health | Dichotomous | 2 or more independent raters | Inter-rater reliability = 97% | Not replicable | Partially reported – All studies (Authors reported number and ratio of missed checklist items for each study) | Vague |
| Dai^S3^ | 2020 | BMC Medical Research Methodology | Multi-level | 2 or more raters (Independence not stated) | N/A | Not replicable | Not reported | Inadequate - Explicit |
| Duan^S4^ | 2020 | BMC Complementary Medicine and Therapies | Multi-level | 2 or more independent raters | Average ICC values of three rounds pre-tests = 0.62, 0.79 and 0.83 respectively | Not replicable | Not reported | Inadequate - Explicit |
| Gore^S5^ | 2020 | Journal of Chronic Obstructive Pulmonary Disease | Dichotomous | 2 or more independent raters | Cohen’s k =0.91 (95%CI 0.79–1.03) | Not replicable | Completely reported | No mention |
| Gundogan^S6^ | 2020 | JAAD International | Multi-level | Not reported | N/A | Not replicable | Partially reported - All studies (Authors reported % of adequately reported items for each study) | Inadequate - Implicit |
| Hogan^S7^ | 2020 | American Journal of Clinical Pathology | Dichotomous | 2 or more independent raters | Cohen’s k = 0.88 (95%CI 0.87-0.89) | Not replicable | Partially reported - Some studies (Authors reported overall adherence (%) and items with < 20% reporting for 5 studies) | Inadequate - Explicit |
| Hou^S8^ | 2020 | Evidence-Based Complementary and Alternative Medicine | Multi-level | 2 or more independent raters | Cohen’s k = 0.93 | Not replicable | Completely reported | Inadequate - Explicit |
| Huang^S9^ | 2020 | Frontiers in Aging Neuroscience | Multi-level | 2 or more independent raters | Not reported | Not replicable | Completely reported | No mention |
| Huang^S10^ | 2020 | Journal of Pain Research | Multi-level | 2 or more independent raters | Not reported | Not replicable | Partially reported - All studies (Overall PRISMA score reported for each individual study) | Inadequate - Implicit |
| Li^S11^ | 2020 | Evidence-Based Complementary and Alternative Medicine | Dichotomous | 2 or more independent raters | Not reported | Not replicable | Completely reported | Inadequate - Explicit |
| Li^S12^ | 2020 | Journal of Dentistry | Dichotomous | 2 or more independent raters | Not reported | Not replicable | Not reported | Inadequate - Explicit |
| Lyu^S13^ | 2020 | Gastroenterology Research and Practice | Multi-level | 2 or more independent raters | Not reported | Not replicable | Completely reported | Inadequate - Explicit |
| Manouchehri^S14^ | 2020 | Iranian Journal of Nursing and Midwifery Research | Not reported | 2 or more independent raters | Not reported | Not replicable | Not reported | Inadequate - Implicit |
| Mazhar^S15^ | 2020 | International Journal of Infectious Diseases | Dichotomous | 1 rater with validation from a second rater | N/A | Partially replicable | Completely reported | Inadequate - Implicit |
| Ngah^S16^ | 2020 | Vaccines (Basel) | Dichotomous | 2 or more independent raters | Not reported | Not replicable | Not reported | Inadequate - Explicit |
| Rainkie^S17^ | 2020 | PLoS One | Dichotomous | 2 or more independent raters | Cohen’s k =  0.82 (PRISMA-P)  0.43 (PRISMA)  % agreement =  92% (PRISMA-P) 91% (PRISMA) | Not replicable | Partially reported - All studies (Authors reported total PRISMA and PRISMA-P scores for each study) | Inadequate - Explicit |
| Suchá^S18^ | 2020 | Radiology: Cardiothoracic Imaging | Dichotomous | 2 or more independent raters | Not reported | Not replicable | Completely reported | Inadequate - Implicit |
| Tian^S19^ | 2020 | Chinese Medicine | Dichotomous | 2 or more independent raters | Not reported | Not replicable | Not reported | Inadequate - Implicit |
| Zhang^S20^ | 2020 | Chinese Acupuncture & Moxibustion | Not reported | 2 or more independent raters | Not reported | Not replicable | Not reported | Inadequate - Explicit |
| Zhu^S21^ | 2020 | International Immunopharmacology | Multi-level | 2 or more independent raters | Not reported | Not replicable | Completely reported | Inadequate - Explicit |
| Adams^S22^ | 2021 | BMJ Open | Multi-level | 2 or more independent raters | ICC= 0.83  (95% CI:0.75 to 0.88) | Fully replicable | Not reported | Vague |
| Adobes Martin^S23^ | 2021 | BMC Medical Research Methodology | Multi-level | 2 or more independent raters | Not reported | Fully replicable | Completely reported | Inadequate - Implicit |
| Almeheyawi^S24^ | 2021 | Journal of Foot and Ankle Research | Dichotomous | 1 rater with validation from a second rater | N/A | Not replicable | Completely reported | No mention |
| Bachelet^S25^ | 2021 | BMC Medical Research Methodology | Other (Each item was measured as a binary outcome or with three ordinal categories) | 2 or more independent raters | 80% concordance | Partially replicable | Completely reported | Inadequate - Explicit |
| Bae^S26^ | 2021 | Complementary Therapies in Clinical Practice | Not reported | 2 or more independent raters | Not reported | Not replicable | Not reported | Inadequate - Explicit |
| Barhli^S27^ | 2021 | Critical Reviews in Oncology / Hematology | Not reported | Not reported | N/A | Not replicable | Not reported | Inadequate - Explicit |
| Beneki^S28^ | 2021 | Journal of Thrombosis and Thrombolysis | Dichotomous | Not reported | N/A | Not replicable | Not reported | Inadequate - Explicit |
| Berton^S29^ | 2021 | Journal of Clinical Medicine | Dichotomous | 2 or more independent raters | % agreement = 99% | Not replicable | Partially reported - All studies (Authors reported number and ratio of missed checklist items for each study) | Vague |
| Besag^S30^ | 2021 | Journal of Psychopharmacology | Multi-level | 2 or more independent raters | Not reported | Not replicable | Completely reported | No mention |
| Bruno^S31^ | 2021 | American Journal of Obstetrics & Gynecology MFM | Dichotomous | 2 or more raters (independence not stated) | N/A | Not replicable | Not reported | Inadequate - Implicit |
| Burton^S32^ | 2021 | Cochrane Database of Systematic Reviews | Multi-level | 2 or more independent raters | Not reported | Not replicable | Completely reported | Inadequate - Explicit |
| Canagarajah^S33^ | 2021 | European Archives of Oto-Rhino-Laryngology | Dichotomous | 1 rater with validation from a second rater | N/A | Not replicable | Not reported | No mention |
| Cao^S34^ | 2021 | Journal of Clinical Epidemiology | Multi-level | 2 or more independent raters | Cohen’s k > 0.68 (in 24/27 items) | Not replicable | Not reported | Vague |
| Chair^S35^ | 2021 | International Journal of Environmental Research and Public Health | Dichotomous | 1 rater with validation from a second rater | N/A | Not replicable | Completely reported | No mention |
| Cho^S36^ | 2021 | Healthcare (Basel) | Dichotomous | 2 or more independent raters | Not reported | Not replicable | Completely reported | Mixed |
| Du^S37^ | 2021 | Thoracic Cancer | Multi-level | 2 or more raters (independence not stated) | N/A | Not replicable | Not reported | Inadequate - Explicit |
| Duan^S38^ | 2021 | The American Journal of Chinese Medicine | Multi-level | 2 or more independent raters | Average ICC values of two rounds of pre-tests were 0.74 and 0.78 (2 articles/round) | Fully replicable | Not reported | Inadequate - Explicit |
| Eliya^S39^ | 2021 | Journal of the American Heart Association | Dichotomous | 2 or more independent raters | Not reported | Partially replicable | Not reported | Vague |
| Escobar-Viera^S40^ | 2021 | Internet Interventions | Dichotomous | 1 rater with validation from a second rater | N/A | Not replicable | Completely reported | No mention |
| Garnier^S41^ | 2021 | Neuro-Oncology Practice | Multi-level | 2 or more independent raters | Median % agreement = 77% (Range 61% -98%) | Not replicable | Partially reported - All studies (P Authors reported total adherence scores for each study) | Inadequate - Explicit |
| Hu^S42^ | 2021 | Journal of Advanced Nursing | Multi-level | 2 or more independent raters | Not reported | Not replicable | Completely reported | No mention |
| Huang^S43^ | 2021 | Pain Research and Management | Multi-level | 2 or more independent raters | Not reported | Not replicable | Completely reported | Vague |
| Jacobsen^S44^ | 2021 | British Journal of Anaesthesia | Multi-level | 2 or more independent raters | Not reported | Not replicable | Partially reported - All studies (Proportion of PRISMA criteria met for each individual study) | Vague |
| Jamnani^S45^ | 2021 | Medical Journal of the Islamic Republic of Iran | Multi-level | 2 or more independent raters | Not reported | Not replicable | Partially reported - Some studies (The authors reported complete ratings for 2 studies as an example) | No mention |
| Jumah^S46^ | 2021 | Stroke | Not reported | 2 or more independent raters | Not reported | Not replicable | Not reported | Inadequate - Explicit |
| Kennedy^S47^ | 2021 | Haemophilia | Dichotomous | 1 rater | N/A | Not replicable | Completely reported | No mention |
| Kim^S48^ | 2021 | Harm Reduction Journal | Multi-level | 2 or more independent raters | Not reported | Not replicable | Completely reported | Vague |
| Knippschild^S49^ | 2021 | BMJ Open | Not reported | 2 or more independent raters | Authors provided Cohen’s k for each item | Not replicable | Not reported | Inadequate - Implicit |
| Kshirsagar^S50^ | 2021 | Journal of Oral and Maxillofacial Surgery | Dichotomous | Other (4 assessed by all authors together and each author assessed 19 articles individually) | N/A | Not replicable | Not reported | Inadequate - Implicit |
| Li^S51^ | 2021 | Annals of Palliative Medicine | Multi-level | 2 or more independent raters | Cohen’s k =0.89 | Not replicable | Completely reported | Inadequate - Implicit |
| Li^S52^ | 2021 | Evidence-Based Complementary and Alternative Medicine | Dichotomous | 2 or more independent raters | Not reported | Not replicable | Completely reported | Inadequate - Explicit |
| Li^S53^ | 2021 | Expert Review of Molecular Diagnostics | Multi-level | 2 or more independent raters | Not reported | Not replicable | Not reported | No mention |
| Li^S54^ | 2021 | Journal of Clinical Epidemiology | Multi-level | 2 or more independent raters | Not reported | Not replicable | Completely reported | No mention |
| Liang^S55^ | 2021 | Chinese Medicine | Multi-level | 2 or more independent raters | Not reported | Not replicable | Completely reported | No mention |
| Liu^S56^ | 2021 | Journal of Pain Research | Dichotomous | 2 or more independent raters | Authors reported Cohen's k for each item | Not replicable | Completely reported | Vague |
| Liu^S57^ | 2021 | Obesity Reviews | Multi-level | 2 or more independent raters | Not reported | Fully replicable | Not reported | Vague |
| Lu^S58^ | 2021 | Phytomedicine | Multi-level | 2 or more independent raters | Not reported | Not replicable | Completely reported | Inadequate - Implicit |
| Malone^S59^ | 2021 | Cancer Medicine | Dichotomous | 2 or more independent raters | Not reported | Not replicable | Not reported | Mixed |
| McGrath^S60^ | 2021 | Journal of Pediatric Urology | Not reported | Not reported | N/A | Not replicable | Not reported | Inadequate - Explicit |
| Morand^S61^ | 2021 | Archives de Pédiatrie | Not reported | Not reported | N/A | Not replicable | Not reported | Adequate |
| Nascimento^S62^ | 2021 | Brazilian Journal of Physical Therapy | Dichotomous | 2 or more independent raters | Authors reported Cohen’s k for each item | Not replicable | Not reported | Inadequate - Explicit |
| Prager^S63^ | 2021 | BMJ Evidence-Based Medicine | Dichotomous | 2 or more independent raters | Cohen’s k =0.59 | Partially replicable | Not reported | Vague |
| Prins^S64^ | 2021 | Archives of Disease in Childhood | Dichotomous | 2 or more independent raters | Cohen’s k = 0.91 (Range 0.60–1.00) | Not replicable | Not reported | Inadequate - Implicit |
| Qin^S65^ | 2021 | European Journal of Orthodontics | Dichotomous | 2 or more independent raters | Not reported | Not replicable | Not reported | Inadequate - Implicit |
| Rod^S66^ | 2021 | Accident Analysis and Prevention | Dichotomous | 1 rater with validation from a second rater | N/A | Not replicable | Completely reported | No mention |
| Shi^S67^ | 2021 | Systematic Reviews | Multi-level | 2 or more independent raters | Not reported | Not replicable | Partially reported - All studies (Authors reported total PRISMA scores for each study) | Inadequate - Implicit |
| Sun^S68^ | 2021 | Nursing Open | Multi-level | 2 or more independent raters | Cohen’s k = 0.76 | Not replicable | Not reported | Inadequate - Explicit |
| Tian^S69^ | 2021 | Evidence-Based Complementary and Alternative Medicine | Multi-level | 2 or more independent raters | Not reported | Not replicable | Completely reported | Inadequate - Implicit |
| Veroniki^S70^ | 2021 | Systematic Reviews | Dichotomous | Not reported | N/A | Not replicable | Not reported | Vague |
| Wenhui^S71,b^ | 2021 | Diabetes Research and Clinical Practice | Multi-level | 2 or more independent raters | Not reported | Not replicable | Not reported | No mention |
| Wright^S72^ | 2021 | Clinical Medicine & Research | Dichotomous | 2 or more independent raters | Not reported | Not replicable | Not reported | Inadequate - Explicit |
| Yang^S73^ | 2021 | International Journal of General Medicine | Dichotomous | 2 or more independent raters | Not reported | Not replicable | Completely reported | Inadequate - Explicit |
| Yin^S74^ | 2021 | PLoS One | Dichotomous | 2 or more independent raters | Not reported | Not replicable | Not reported | Inadequate - Explicit |
| Yuan^S75^ | 2021 | Evidence-Based Complementary and Alternative Medicine | Multi-level | 2 or more independent raters | Not reported | Not replicable | Completely reported | Vague |
| Zhang^S76^ | 2021 | Asian Pacific Journal of Clinical Nutrition | Multi-level | 2 or more independent raters | Cohen’s k =0.93 | Not replicable | Not reported | Inadequate - Explicit |
| Zhang^S77^ | 2021 | Chinese Medicine | Dichotomous | 2 or more independent raters | Not reported | Fully replicable | Not reported | Inadequate - Implicit |
| Zhang^S78^ | 2021 | Frontiers in Pharmacology | Multi-level | 2 or more independent raters | Not reported | Not replicable | Completely reported | Inadequate - Implicit |
| Zhang^S79^ | 2021 | Journal of Clinical Epidemiology | Dichotomous | 2 or more independent raters | Authors reported interrater agreement for each item | Not replicable | Not reported | Inadequate - Implicit |
| Zheng^S80^ | 2021 | Annals of Translational Medicine | Not reported | 2 or more independent raters | Not reported | Not replicable | Not reported | Inadequate - Explicit |
| Zheng^S81^ | 2021 | Frontiers in Medicine | Multi-level | 2 or more independent raters | Not reported | Not replicable | Not reported | Inadequate - Explicit |
| Al-Abedalla^S82^ | 2022 | JDR Clinical & Translational Research | Dichotomous | 2 or more independent raters | Not reported | Not replicable | Partially reported - All studies (Authors reported overall CONSORT score for each study) | Inadequate - Explicit |
| Alshahwani^S83^ | 2022 | Journal of Surgical Research | Multi-level | 2 or more independent raters | Not reported | Not replicable | Partially reported - All studies (Authors reported total percentage of reported items for PRISMA for each study) | Adequate |
| Astur^S84^ | 2022 | einstein | Multi-level | 2 or more independent raters | Not reported | Not replicable | Not reported | Vague |
| Batioja^S85^ | 2022 | Journal of Pediatric Orthopaedics | Multi-level | 2 or more independent raters | Not reported | Not replicable | Not reported | Inadequate - Explicit |
| Bole^S86^ | 2022 | The Journal of Sexual Medicine | Not reported | 2 or more raters (independence not stated) | N/A | Not replicable | Not reported | Inadequate - Implicit |
| Bonetti^S87^ | 2022 | Research in Social and Administrative Pharmacy | Dichotomous | 2 or more independent raters | Not reported | Not replicable | Not reported | No mention |
| Cervantes^S88^ | 2022 | Psychiatric Services | Not reported | 1 rater | N/A | Not replicable | Partially reported - All studies (Authors reported total STROBE adherence (%) for each study) | No mention |
| Chen^S89^ | 2022 | Journal of Ginseng Research | Dichotomous | 2 or more independent raters | Not reported | Not replicable | Completely reported | Vague |
| Cindro^S90^ | 2022 | BMJ Open | Dichotomous | 2 or more independent raters | Cohen’s k > 0.60 for all items | Not replicable | Not reported | Inadequate - Explicit |
| de Lucena Alves^S91^ | 2022 | Clinical Implant Dentistry and Related Research | Dichotomous | 2 or more independent raters | Not reported | Not replicable | Not reported | Mixed |
| Douglas^S92^ | 2022 | BMJ Evidence-Based Medicine | Other (Both dichotomous and multilevel methods were used) | 2 or more independent raters | Not reported | Not replicable | Not reported | Inadequate - Explicit |
| Feng^S93^ | 2022 | Clinical and Translational Gastroenterology | Multi-level | 2 or more independent raters | Not reported | Partially replicable | Not reported | Inadequate - Implicit |
| Fernández-Pires^S94^ | 2022 | The American Journal of Occupational Therapy | Dichotomous | 2 or more independent raters | Cohen’s k ≥ 0.80 (Random sample of 34 articles) | Not replicable | Not reported | Mixed |
| Frank^S95^ | 2022 | Journal of Magnetic Resonance Imaging | Dichotomous | 2 or more independent raters | Not reported | Fully replicable | Not reported | No mention |
| Gebran^S96^ | 2022 | Journal of the American College of Surgeons | Dichotomous | 2 or more independent raters | % agreement = 84% | Not replicable | Not reported | Inadequate - Explicit |
| Gupta^S97^ | 2022 | Perspectives in Clinical Research | Dichotomous | Not reported | N/A | Not replicable | Not reported | Vague |
| Helliwell^S98^ | 2022 | Health Science Reports | Not reported | 2 or more independent raters | Not reported | Not replicable | Not reported | Vague |
| Huang^S99^ | 2022 | Frontiers in Public Health | Multi-level | 2 or more independent raters | Not reported | Not replicable | Completely reported | Inadequate - Implicit |
| Imran^S100^ | 2022 | Journal of Clinical Epidemiology | Multi-level | 1 rater with validation from a second rater | N/A | Fully replicable | Completely reported | Inadequate - Explicit |
| Jung^S101^ | 2022 | Clinical Microbiology and Infection | Not reported | Not reported |  | Not replicable | Partially reported - All studies (Authors reported total CONSORT score for each study) | Adequate |
| Kanukula^S102^ | 2022 | BMJ Evidence-Based Medicine | Dichotomous | 2 or more independent raters | Not reported | Not replicable | Partially reported - All studies (Authors reported PRISMA index score for each study) | Mixed |
| Kazi^S103^ | 2022 | Journal of Magnetic Resonance Imaging | Dichotomous | 2 or more independent raters | Cohen’s k =  0.65 to 0.70 (STARD);  0.71 to 0.76 (STARD for Abstracts) | Fully replicable | Not reported | Vague |
| Khachfe^S104^ | 2022 | The American Journal of Surgery | Dichotomous | 2 or more independent raters | % agreement =  91% (STROBE)  92% (RECORD) | Not replicable | Partially reported - All studies (Authors reported total reporting guideline scores for each study) | Adequate |
| Khan^S105^ | 2022 | Multiple Sclerosis and Related Disorders | Dichotomous | 2 or more independent raters | Not reported | Not replicable | Not reported | Inadequate - Explicit |
| Kim^S106^ | 2022 | Frontiers in Medicine | Dichotomous | 2 or more independent raters | Not reported | Not replicable | Completely reported | No mention |
| Labiste^S107^ | 2022 | Skeletal Radiology | Not reported | 2 or more independent raters | Not reported | Not replicable | Partially reported - All studies (Authors reported average STARD adherence for each study) | No mention |
| Li^S108^ | 2022 | Evidence-Based Complementary and Alternative Medicine | Multi-level | 2 or more independent raters | Not reported | Not replicable | Completely reported | Inadequate - Implicit |
| Love^S109^ | 2022 | Critical Reviews in Oncology / Hematology | Multi-level | 2 or more independent raters | Not reported | Not replicable | Partially reported - All studies (Authors reported total PRISMA score for each study) | Inadequate - Explicit |
| Lu^S110^ | 2022 | Journal of Integrative Medicine | Multi-level | 2 or more independent raters | Not reported | Not replicable | Completely reported | Inadequate - Explicit |
| McCall^S111^ | 2022 | Journal of Clinical Epidemiology | Multi-level | 1 rater with validation from a second rater | N/A | Fully replicable | Completely reported | Inadequate - Explicit |
| Mc Cord^S112^ | 2022 | Journal of Clinical Epidemiology | Multi-level | 2 or more independent raters | Not reported | Fully replicable | Completely reported | Inadequate - Explicit |
| McErlean^S113^ | 2022 | Irish Journal of Medical Science | Not reported | 2 or more independent raters | Not reported | Not replicable | Partially reported - All studies (Authors reported number of adhered CONSORT items for each study) | Mixed |
| Menne^S114^ | 2022 | Journal of Periodontology | Multi-level | Other (Two reviewers extracted data in duplicate and independently for 30 abstracts. Differences were discussed until agreement reached 80% and thereafter data extraction was done by one reviewer.) | N/A | Fully replicable | Not reported | Inadequate - Explicit |
| Newman^S115^ | 2022 | Diabetes Research and Clinical Practice | Dichotomous | 2 or more independent raters | Not reported | Not replicable | Not reported | Vague |
| Park^S116^ | 2022 | Korean Journal of Radiology | Not reported | 2 or more raters (Independence not stated) | N/A | Not replicable | Partially reported - Some studies (The authors report articles that did not adhere to particular PRISMA guideline items) | Inadequate - Implicit |
| Peña^S117^ | 2022 | Urology | Multi-level | 2 or more independent raters | Not reported | Not replicable | Partially reported - All studies (Authors reported total PRISMA score for each study) | Mixed |
| Pfannenstiel^S118^ | 2022 | The International Journal of Oral & Maxillofacial Implants | Multi-level | 2 or more raters (Independence not stated) | N/A | Fully replicable | Not reported | Inadequate - Explicit |
| Ruan^S119^ | 2022 | International Journal of General Medicine | Dichotomous | 2 or more raters (Independence not stated) | N/A | Not replicable | Not reported | Inadequate - Explicit |
| Shi^S120^ | 2022 | Cardiology Research and Practice | Dichotomous | 2 or more independent raters | Not reported | Not replicable | Completely reported | Vague |
| Shi^S121^ | 2022 | Drug Design, Development and Therapy | Multi-level | 2 or more independent raters | Not reported | Not replicable | Completely reported | Inadequate - Implicit |
| Shin^S122^ | 2022 | International Journal of Environmental Research and Public Health | Multi-level | 2 or more independent raters | Not reported | Not replicable | Completely reported | No mention |
| Siew^S123^ | 2022 | Psychoneuroendocrinology | Dichotomous | 1 rater | N/A | Not replicable | Partially reported - All studies (Authors provided completeness of reporting score (COR) for each study) | No mention |
| Streck^S124^ | 2022 | Nicotine and Tobacco Research | Multi-level | 2 or more raters (Independence not stated) | N/A | Not replicable | Partially reported - All studies (Authors reported PRISMA percent complete for each study) | Inadequate - Explicit |
| Stunnenberg^S125^ | 2022 | Neurology | Dichotomous | Not reported | N/A | Not replicable | Not reported | Inadequate - Explicit |
| Tanner^S126^ | 2022 | Drug and Alcohol Dependence | Multi-level | 2 or more raters (Independence not stated) | N/A | Not replicable | Partially reported - All studies (Authors reported PRISMA % complete for each study) | Vague |
| Torgerson^S127^ | 2022 | International Journal of Pediatric Otorhinolaryngology | Multi-level | 2 or more independent raters | Not reported | Not replicable | Partially reported - All studies (Authors reported total PRISMA score for each study) | Mixed |
| Warrier^S128^ | 2022 | Perspectives in Clinical Research | Not reported | Not reported | N/A | Not replicable | Not reported | Inadequate - Explicit |
| Yang^S129^ | 2022 | Evidence-Based Complementary and Alternative Medicine | Multi-level | 2 or more independent raters | Authors reported Cohen’s k coefficient for each item | Not replicable | Completely reported | Inadequate - Implicit |
| Yang^S130^ | 2022 | Journal of Ethnopharmacology | Dichotomous | 2 or more independent raters | Not reported | Not replicable | Completely reported | Inadequate - Explicit |
| Yao^S131^ | 2022 | Journal of Integrative Medicine | Multi-level | 2 or more independent raters | Cohen’s k = 0.91 | Not replicable | Completely reported | No mention |
| Yin^S132^ | 2022 | Frontiers in Physiology | Multi-level | 2 or more independent raters | Not reported | Not replicable | Completely reported | No mention |
| Yin^S133^ | 2022 | International Journal of Infectious Diseases | Multi-level | 2 or more independent raters | Not reported | Not replicable | Not reported | Inadequate - Explicit |
| Yuniar^S134^ | 2022 | Vaccines (Basel) | Dichotomous | 2 or more independent raters | Cohen's k = 0.79 | Not replicable | Not reported | Adequate |
| Zhang^S135^ | 2022 | Human Vaccines & Immunotherapeutics | Not reported | 2 or more independent raters | Authors reported Cohen’s k for each item | Not replicable | Not reported | Adequate |
| Zhou^S136^ | 2022 | Therapeutic Advances in Gastroenterology | Not reported | 1 rater with validation from a second rater | N/A | Not replicable | Not reported | Inadequate - Implicit |
| Ziemann^S137^ | 2022 | BMC Medical Research Methodology | Dichotomous | 1 rater with validation from a second rater | N/A | Fully replicable | Partially reported - All studies (Authors reported % STROBE completion for each study) | Inadequate - Explicit |
| Anderson^S138^ | Online only | The Journal of Arthroplasty | Dichotomous | 2 or more independent raters | Not reported | Fully replicable | Not reported | Inadequate - Explicit |
| Bonafiglia^S139^ | Online only | Journal of Sport and Health Science | Dichotomous | 2 or more independent raters | Not reported | Not replicable | Completely reported | No mention |
| Brito-Suárez^S140^ | Online only | Pediatric Hematology and Oncology | Dichotomous | 2 or more independent raters | Not reported | Not replicable | Completely reported | No mention |
| Cremades-Martínez^S141^ | Online only | Microbiology Spectrum | Dichotomous | 2 or more independent raters | Not reported | Not replicable | Completely reported | Vague |
| Dhillon^S142^ | Online only | The Laryngoscope | Multi-level | 2 or more independent raters | Not reported | Not replicable | Partially reported - All studies (Authors reported total PRISMA score for each study) | Mixed |
| Garrett^S143^ | Online only | BMJ Evidence-Based Medicine | Multi-level | 2 or more independent raters | Not reported | Not replicable | Partially reported - All studies (Authors provided percent PRISMA complete for each study) | Inadequate - Explicit |
| Gysling^S144^ | Online only | Seminars in Thoracic and Cardiovascular Surgery | Dichotomous | 2 or more independent raters | Authors reported Cohen’s k for each item | Not replicable | Not reported | Inadequate - Explicit |
| Innocenti^S145^ | Online only | Archives of Physical Medicine and Rehabilitation | Dichotomous | 2 or more independent raters | Cohen’s k = 0.83 | Not replicable | Not reported | Inadequate - Explicit |
| Kim^S146^ | Online only | Academic Radiology | Dichotomous | 2 or more independent raters | Cohen’s k = 0.75 | Not replicable | Not reported | Inadequate - Implicit |
| Snider^S147^ | Online only | Clinical Breast Cancer | Multi-level | 2 or more independent raters | Not reported | Not replicable | Partially reported - All studies (Authors reported PRISMA % complete for each study) | Mixed |
| Tosatto^S148^ | Online only | Restorative Neurology and Neuroscience | Dichotomous | 2 or more independent raters | Cohen’s k = 0.88 | Not replicable | Not reported | Inadequate - Implicit |

^a^Year of publication based on when data were extracted.

^b^Contacted corresponding authors 3 times asking for supplementary file and did not receive a response. We also contacted the corresponding journal manager, who then contacted the authors as well, and never heard back from them. The outcomes are based on data that were available to us.
